# Proteoform-resolved neoGFAP^™^ as a diagnostic and prognostic biomarker across the TBI–MCI–AD continuum in Veterans

**DOI:** 10.64898/2026.09.01.26361845

**Authors:** William E. Haskins, Kevin K. Wang, Guangzheng Cai, Khadija Boukholda, Eman Elbayoumi, Ruchi Bajpai, Devin Jackson, Katie Tehas, Kristy Radeker, Anthony DeLizza, Caroline Popper, Martin Kiendl, Sigrun Badrnya, Markus Miholits, Stefan Jellbauer, Todd Kilbaugh, Franklin Okumu, Ava Puccio, Raquel C. Gardner, Geoff Manley, John B. Williamson, Abigail B. Waters, Gail Ge Li, Elaine R. Peskind

## Abstract

Service members with traumatic brain injury are at approximately two- to four-fold higher risk of Alzheimer’s disease or related dementias than those without such an injury, with risk increasing with injury severity. The amyloid/tau/neurodegeneration biomarker framework treats amyloid, tau, and neurodegeneration as independent axes but omits astroglial injury, despite evidence that reactive astrogliosis (indexed by glial fibrillary acidic protein, GFAP) must be elevated for cognitive decline to occur in amyloid-positive individuals. Total GFAP immunoassays aggregate intact protein with multiple calpain- and caspase-cleaved proteoforms, blurring the biological signal. We compared a calpain-cleaved GFAP neoepitope, the glial fibrillary acidic protein neoepitope (neoGFAP™), against total GFAP across the full traumatic-brain-injury–mild-cognitive-impairment–Alzheimer’s-disease continuum in Veterans using a two-stage plasma-to-cerebrospinal-fluid biomarker approach. A plasma triage gate combining phosphorylated tau 217 and amyloid beta 42 was applied to 367 unique subjects; a cerebrospinal-fluid benchmarking cohort of 57 subjects (controls, chronic blast traumatic brain injury, mild cognitive impairment, and Alzheimer’s disease) received head-to-head neoGFAP™ and total GFAP measurement. In the whole benchmarking cohort, neoGFAP™ discriminated mild cognitive impairment plus Alzheimer’s disease from non-Alzheimer subjects with an area under the receiver-operating-characteristic curve of 0.81 versus 0.73 for total GFAP, a trend-level advantage that did not reach nominal significance. Within the gate-positive, amyloid-committed subset of 23 subjects, neoGFAP™ dominance became significant by McNemar’s exact test (six discordant subjects favoured neoGFAP™, none the reverse). Across diagnostic contrasts, neoGFAP™ outperformed total GFAP for Alzheimer’s disease versus control and, importantly for Veterans, for mild cognitive impairment versus chronic blast-exposed Veterans without cognitive impairment. In chronic blast injury, neoGFAP™ was paradoxically depleted relative to controls, consistent with tissue sequestration of aggregated proteoform fragments. Unbiased proteomic profiling confirmed coordinated elevation across astrocytic, neuronal, mitochondrial, and microglial compartments. An exploratory subject-level reclassification improved accuracy from 71.1 percent using plasma alone to 79.5 percent with added cerebrospinal-fluid markers and age. In a same-cohort ProQuantum™ replication (n=57), CSF neoGFAP™ preserved its discrimination advantage over total GFAP for MCI+AD versus non-AD (AUROC 0.76 vs 0.72; cross-platform Spearman ρ=0.84), while plasma neoGFAP™ achieved AUROC 0.90, comparable to pTau217 (0.92) and exceeding Aβ42/40 (0.84). In this small sample, neoGFAP™ is a superior proteoform-resolved diagnostic and prognostic biomarker across the continuum and supports adding an astroglial-proteoform axis to amyloid/tau/neurodegeneration biomarker frameworks in high-risk populations.

**Abbreviated summary:** Haskins and colleagues report that a calpain-cleaved GFAP neoepitope (neoGFAP™) outperforms total GFAP for diagnostic and prognostic classification across the traumatic-brain-injury–mild-cognitive-impairment–Alzheimer’s-disease continuum in Veterans, supporting the addition of an astroglial-proteoform axis to amyloid/tau/neurodegeneration biomarker frameworks in high-risk populations.

## Introduction

Traumatic brain injury (TBI) is a recognized risk factor for Alzheimer’s disease (AD) and AD-related dementias. In a propensity-matched cohort of 178,779 United States Veterans, a TBI diagnosis was associated with an adjusted hazard ratio for dementia of 2.36 (95% CI 2.10–2.66) after mild TBI without loss of consciousness, rising to 3.77 (95% CI 3.63–3.91) after moderate-to-severe TBI, indicating that the magnitude of the increase depends on injury severity.^19^ TBI induces a wave of extracellular glutamate that exceeds the buffering potential of astrocytic GLT-1; calcium influx activates calpains, which cleave GFAP—the major astrocytic intermediate filament—at an N-terminal site near residue 60, generating GFAP breakdown products with a lower molecularweight limit of 38 kDa. We refer to these calpain-cleaved 38–40 kDa proteoforms as neoGFAP™.^2^

Following injury, neoGFAP™ is released into extracellular fluid, cerebrospinal fluid (CSF), and blood with sustained elevation lasting days to decades post-injury and correlating with functional outcome.^3^ In parallel, cerebrospinal fluid and serum profiles of the axonal-injury markers neurofilament light chain and phosphorylated neurofilament heavy chain follow comparable temporal trajectories and are associated with patient outcome after moderate-severe TBI.^4^ Long-term impairment of central-nervous-system debris clearance—through glymphatic, lymphatic, and microglial or phagocytic dysfunction—sustains this elevation and provides a mechanistic bridge from TBI to mild cognitive impairment (MCI) to AD.

The amyloid/tau/neurodegeneration (A/T/N) framework for AD diagnosis and prognosis defines amyloid, tau, and neurodegeneration as independent biomarker axes.^5^ Recent landmark studies show this framework is incomplete: an AD-like pattern of tau accumulation as a function of amyloid beta was observed only in cognitively unimpaired individuals with high GFAP, suggesting that astrocyte reactivity is an upstream gating event linking amyloid beta to tau pathology.^6^ Convergent conclusions were reached using plasma GFAP.^7,8,9^ Critically, these studies all used total-GFAP immunoassays that conflate intact 50 kDa GFAP with multiple calpain- and caspase-generated proteoforms.^10^ If the biologically active discriminant signal resides in a specific proteoform, a proteoform-resolved assay should outperform total GFAP for diagnostic classification.

Here we compare CSF neoGFAP™ with total GFAP across the full TBI–MCI–AD continuum using a two-stage plasma-to-CSF biomarker scheme. Stage 1 is a non-invasive plasma triage gate built from the Lumipulse pTau217 and amyloid beta 42 combination (pTau217 above the 75th-percentile control cutoff and amyloid beta 42 below the 25th-percentile control cutoff), designed to identify subjects enriched for AD-pathway pathology from a single blood draw. Recent work from the DOD-ADNI/ADBI consortium demonstrated that plasma pTau217/Aβ42 accuracy for amyloid-PET status drops sharply in Veterans with a TBI history (from 90% in TBI-negative Veterans to 63% with loss-of-consciousness >5 min), missing over half of amyloid-PET-positive cases in the higher-exposure group,^11^ underscoring the need for a confirmatory step in TBI-exposed populations. Stage 2, applied only within the Stage-1 gate-positive subset, uses neoGFAP™ (CSF in the present benchmarking cohort; plasma in the planned extension) to resolve MCI-like from AD-like astroglial proteoform burden—converting a binary AD-pathway call into a graded severity readout. The two stages are deployed sequentially: Stage 1 first, in plasma, to reduce the population needing invasive sampling, and Stage 2 second, in the smaller gate-positive subset, to grade severity and prioritize. We anchor the findings with unbiased Olink proteomics spanning astrocytic, neuronal, mitochondrial, and microglial compartments. This chronic-phase continuum analysis is submitted in parallel with three acute-phase clinical neoGFAP™ studies (pediatric TBI, CENTER-TBI multinational adult, adult severe TBI) and one preclinical therapeutic-antibody characterization; together they span the injury lifecycle from acute plasma release through chronic neurodegeneration.

## Materials and methods

### Study cohort and samples

This is a cross-sectional, retrospective, single-site observational biomarker study conducted according to STROBE reporting guidance for cohort studies. Archived CSF and plasma samples were obtained under Institutional Review Board approval at the VA Puget Sound Health Care System (ADTBI IRB #1653649) from the VA Northwest Mental Illness Research, Education, and Clinical Center (VA NW MIRECC) Sample and Data Biorepository. The CSF benchmarking cohort for the two-level gating scheme comprised 57 subjects with complete Lumipulse CSF amyloid beta 42/40 and pTau181/tTau plus MSD neoGFAP™ and total GFAP measurements: controls (n = 10), chronic blast TBI (n = 17), MCI (n = 10), and AD (n = 20). The plasma screening cohort (Stage 1) comprised 367 unique subjects (738 plasma sample-rows) with paired amyloid beta 42 and pTau217. Group assignments (Control, TBI, MCI, AD) were taken from the PrimaryDx field recorded in the VA NW MIRECC source workbooks, reflecting the clinical consensus diagnosis assigned at each ADRC consensus conference; no post-hoc relabeling was performed. TBI was restricted to the chronic-blast-exposed deployed subset for all between-group comparisons. Because the source populations were sampled as they exist clinically, age and sex are near-perfectly confounded with diagnostic category (Table 1b); this is addressed explicitly under Limitations.

**Table 1.**
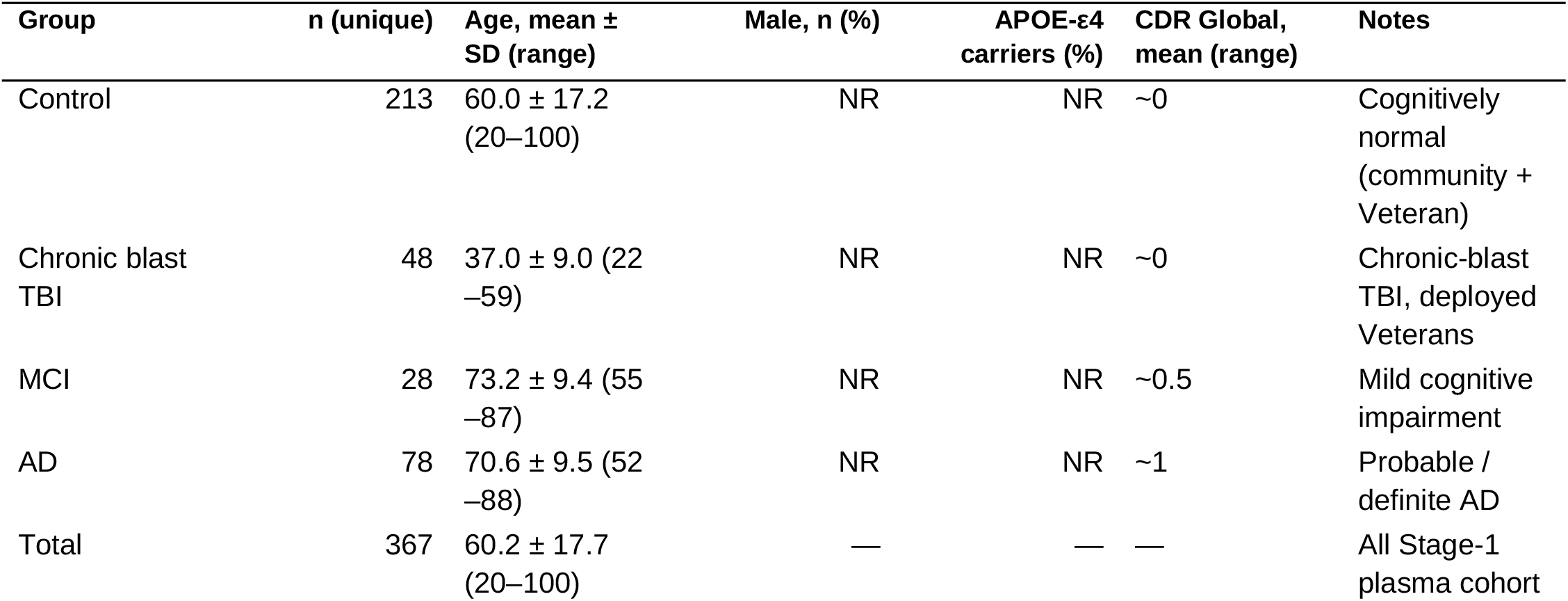
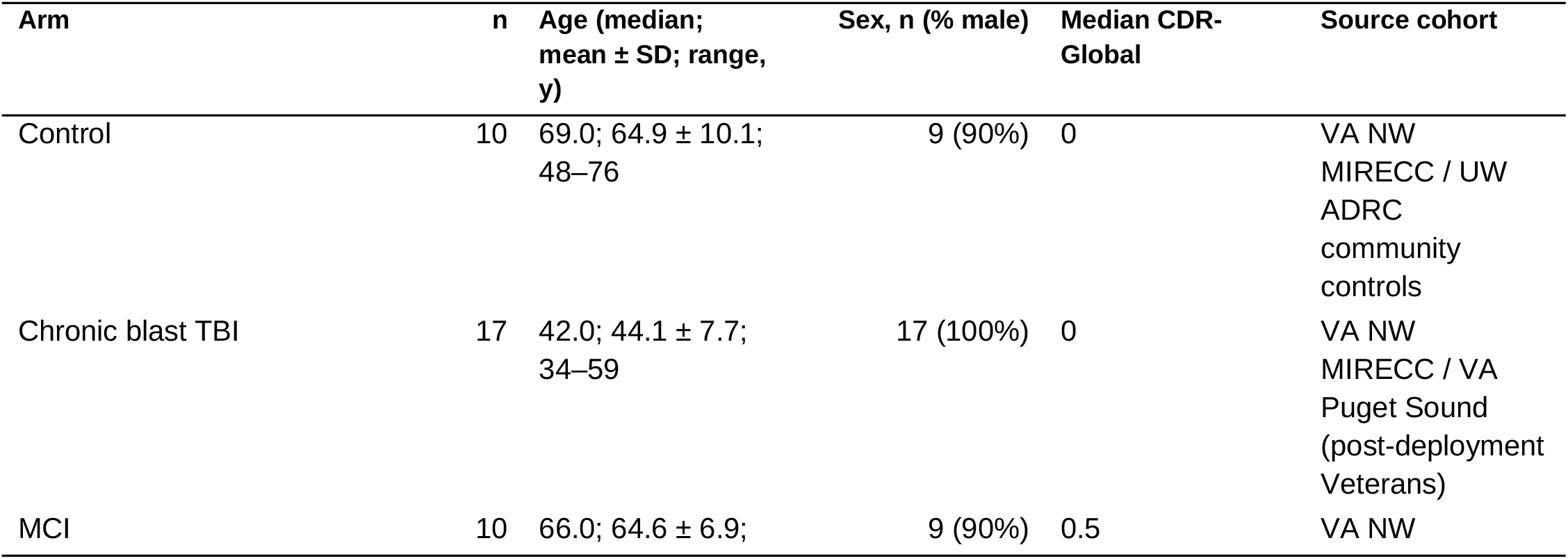
Stage-1 plasma cohort demographic and clinical characteristics (n = 367 unique subjects; 738 plasma sample-rows).

**Table 1b.**
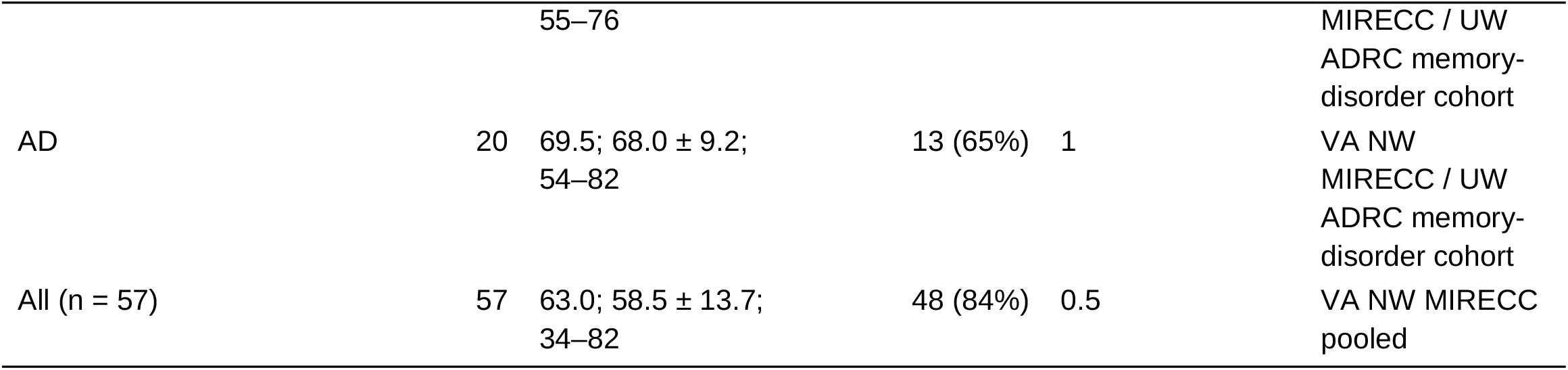
Per-arm demographic breakdown of the CSF benchmarking cohort (n = 57), showing the age/sex/source confound. Age and sex are near-perfectly confounded with diagnostic category: the chronic blast TBI arm (mean 44 y, 100% male) is drawn from a Veteran deployment-injury cohort, whereas the MCI and AD arms (mean 65–68 y, 65–90% male) are drawn from the community memory-disorder cohort. This limitation is discussed in the Limitations section.

### Biomarker assays

All biomarker measurements are tagged by biofluid to remove ambiguity: plasma amyloid beta 40, plasma amyloid beta 42, plasma amyloid beta 42/40, and plasma pTau217 (Lumipulse, plasma); CSF amyloid beta 40/42, CSF pTau181, CSF tTau (Lumipulse, CSF); and CSF neoGFAP™ and CSF total GFAP (Gryphon-MSD assay, CSF, n = 57). Plasma amyloid beta 40, 42, and pTau217 were measured on the Fujirebio Lumipulse G1200 platform; CSF amyloid beta 40/42, pTau181, and tTau were measured on the same platform. CSF neoGFAP™ and total GFAP were measured on the Meso Scale Discovery (MSD) electrochemiluminescence platform using proprietary capture/detection monoclonal antibody pairs and calibrators; these MSD measurements constitute the legacy discovery dataset on which the primary analyses reported here were built, and are reported separately from the ProQuantum replication described below. Plasma-compartment neoGFAP™ and total GFAP were available only for 35 mild/moderate AD subjects and are not yet extended to the other groups; consequently every reclassification analysis uses CSF-compartment GFAP markers.

### ProQuantum replication assays

An independent replication of the astroglial-proteoform measurements was performed on a second platform, chosen so that no single vendor’s chemistry drove the conclusions. ProQuantum is an immuno-PCR platform that couples antibody capture to a quantitative PCR readout; astroglial proteoforms were measured with Invitrogen™ ProQuantum High-Sensitivity Immunoassay Kits on Applied Biosystems™ QuantStudio™ qPCR instruments, using proprietary capture/detection monoclonal antibody pairs and protein calibrators; as in the legacy MSD configuration, Gryphon Bio critical reagents supplied the neoGFAP™ and total GFAP calibrators. The replication used archived, matched aliquots from the same 57 benchmarking subjects, comprising 57 plasma specimens and 57 matched CSF specimens (114 paired plasma and CSF specimens in total), so the ProQuantum panel provides both a same-cohort cross-platform check of the legacy CSF measurements and the first paired plasma measurements of neoGFAP™ and total GFAP in this cohort. Samples were assayed in replicate wells at matrix-appropriate dilutions; analyte concentrations in pg/mL were interpolated from calibrator standard curves and corrected for the sample dilution factor, and the coefficient of variation across replicate wells was retained alongside the cycle-threshold value and concentration for every sample. ProQuantum concentration tables were returned by the assay laboratory keyed only to study subject identifier, sex, and age, with no diagnostic-group field; group assignments were merged locally after the concentration tables were received and locked, and all group-wise comparisons and receiver-operating-characteristic analyses were performed thereafter. Analyte definitions were unchanged between platforms: neoGFAP™ denotes the calpain-cleaved 38–40 kDa GFAP neoepitope and total GFAP denotes the aggregate intact-plus-proteoform GFAP measurand. The legacy MSD discovery analyses and ProQuantum replication analyses were conducted as separate within-platform analyses.

### Two-level gating scheme

Stage-1 plasma cutoffs were derived from the 75th and 95th percentiles of plasma pTau217 and the 25th percentile of plasma amyloid beta 42 in the Lumipulse plasma control pool. The asymmetric percentile choice reflects biological direction: plasma pTau217 rises with AD pathology while amyloid beta 42 falls with amyloid sequestration, so the AD-end tail of each distribution is the pathology-positive tail. Subjects were assigned to Stage 0 (no signal), Stage 1 (borderline—one marker positive), Stage 2 (biomarker positive—both markers at the low cutoff), or Stage 3 (high AD—pTau217 at the high cutoff and amyloid beta 42 at the low cutoff). Stage-2 CSF cutoffs were derived from the 75th percentile of CSF neoGFAP™ and total GFAP in the CSF control reference set (n = 10). The Stage-1/Stage-2 gating scheme is central to the Results: Stage 1 triages in plasma, and Stage 2 applies the proteoform head-to-head only within the gate-positive, amyloid-committed subset.

### Unbiased proteomic analysis

Olink Explore 3072 normalized protein expression (NPX) data were analyzed genome-wide: all 2,908 assays passing quality control were carried forward without curation or a priori restriction. For visualization only, proteins were post-hoc annotated to cell-type compartments (astrocyte, neuronal/axonal, microglia/myeloid, oligodendrocyte/myelin, mitochondria, inflammatory cytokine, endothelial/blood–brain barrier, and others). Five proteoforms/proteins of central interest—CSF neoGFAP™, CSF total GFAP, NEFL, IL6, and PPIF—were flagged in every panel regardless of significance but were not used to filter or weight the analysis. Per-group mean NPX, two-sided Mann–Whitney U p-values against each reference choice, and compartment-stratified significant-protein counts were tabulated across the full proteome.

### Statistical analysis

Group comparisons used the Mann–Whitney U test (one- and two-sided as specified) and the Kruskal–Wallis test. Areas under the receiver-operating-characteristic curve (AUROCs) were compared by paired bootstrap (5,000 iterations) with z-approximation p-values and 95 percent bootstrap confidence intervals. Categorical gate enrichment used Fisher exact tests. Classification dominance between neoGFAP™ and total GFAP on the same subjects used McNemar’s test with exact binomial p-values. Operating points, where reported, used the Youden index to select a threshold. The primary diagnostic analysis comprised five AUROC contrasts (TBI vs healthy, MCI vs healthy, AD vs healthy, AD vs MCI, TBI vs MCI); Benjamini–Hochberg false-discovery-rate (FDR) control at alpha = 0.05 was applied across these five, with Holm-adjusted p-values also reported. Under BH-FDR, four of five primary contrasts remained significant (all except the null MCI-diagnostic contrast). Secondary Olink discovery p-values were not corrected and are reported as nominal. The pre-specified Stage-2-gated McNemar test is a single secondary test with a distinct hypothesis and is not subject to the primary diagnostic-family correction. Analyses used Python with statsmodels for multiple comparisons.

## Results

### Cohort profile and Stage-1 plasma gate

The CSF benchmarking cohort (n = 57) spanned cognitively normal controls, chronic blast TBI, MCI, and AD (Table 1b), while the plasma-screened population contributed 367 subjects stratifiable into Stage 0–3 categories. The Stage-1 plasma gate (Aβ42/pTau217) enriched strongly for AD-pathway pathology: gate-positive rates rose monotonically across clinical groups—control 8.0 percent, TBI 2.1 percent, MCI 35.7 percent, and AD 70.5 percent (Figure 3a) (Supplementary Table S1). The Stage-1 plasma Aβ42/pTau217 ratio discriminated MCI plus AD from controls with an AUROC of 0.93 (95% CI 0.89–0.96; n = 319 labelled subjects), validating Stage-1 stratification and placing the gate at the upper end of published Lumipulse plasma performance (Figure 3b).^12,13^

**Figure 1.**
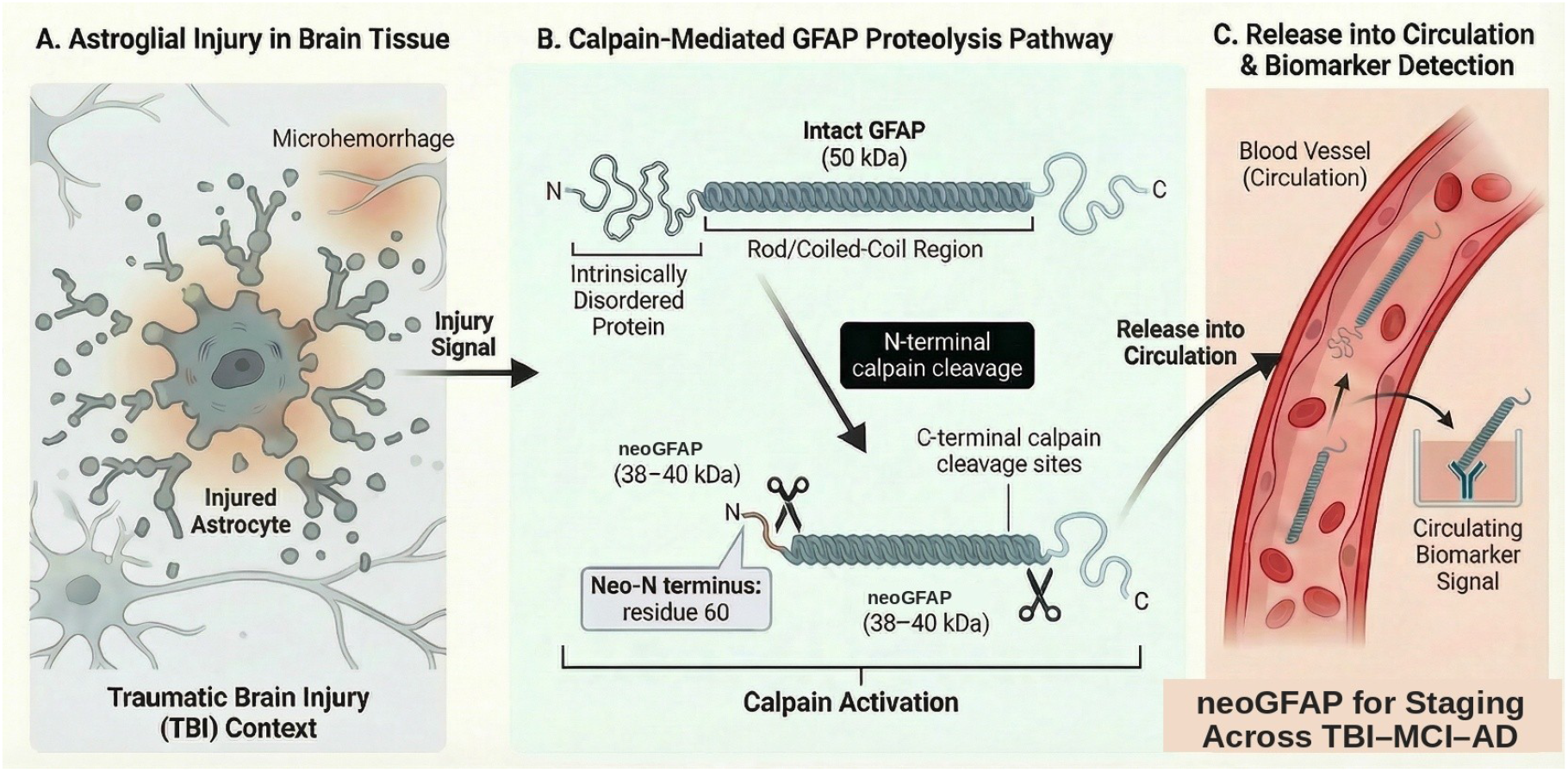
Mechanistic basis of neoGFAP™ generation and release into circulation. (A) Astroglial injury in traumatic brain injury (TBI), showing an injured astrocyte with tissue-damage features such as microhemorrhage that initiate downstream proteolysis. (B) Calpainmediated GFAP proteolysis: N-terminal calpain cleavage exposes a neo-N terminus at residue 60, and additional C-terminal cleavage generates predominantly 38–40 kDa neoGFAP™ fragments. (C) Release of neoGFAP™ proteolytic fragments into the bloodstream, where they can be detected as a circulating biomarker.

**Figure 2.**
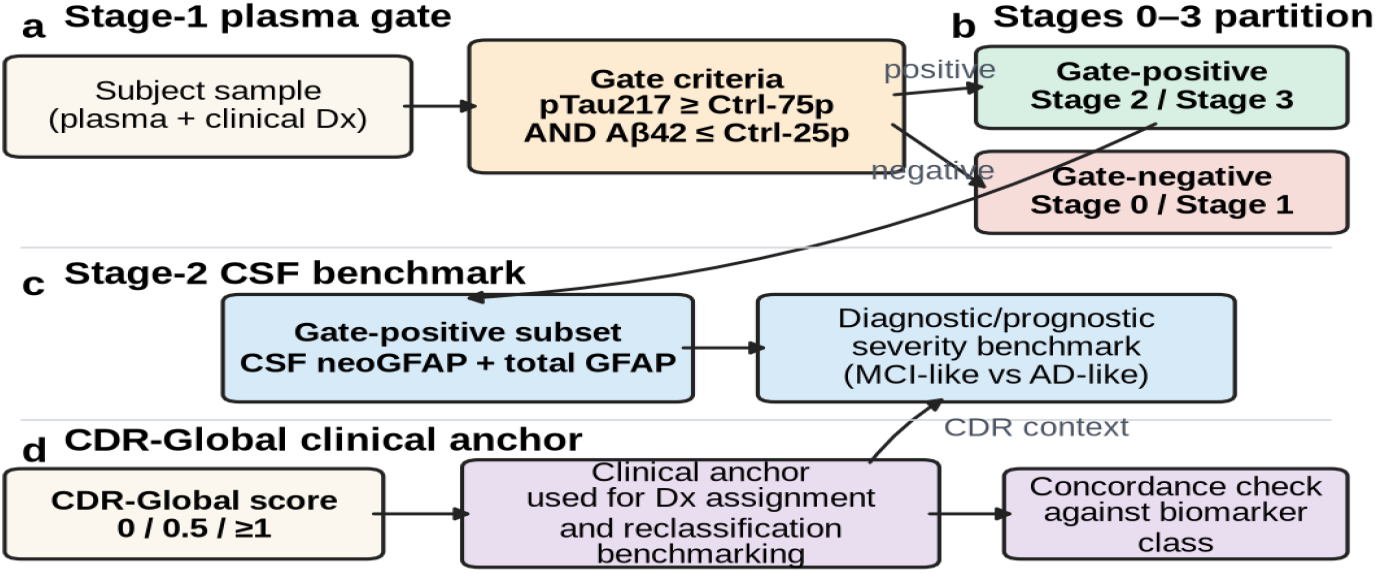
Two-level gating scheme with CDR-Global anchor. The Stage-1 plasma gate (pTau217 ≥ control 75th percentile AND amyloid beta 42 ≤ control 25th percentile) partitions subjects into stages 0–3; the Stage-2 CSF benchmark (neoGFAP™ and total GFAP) is applied only within the Stage-1 gate-positive, amyloid-committed subset.

**Figure 3.**
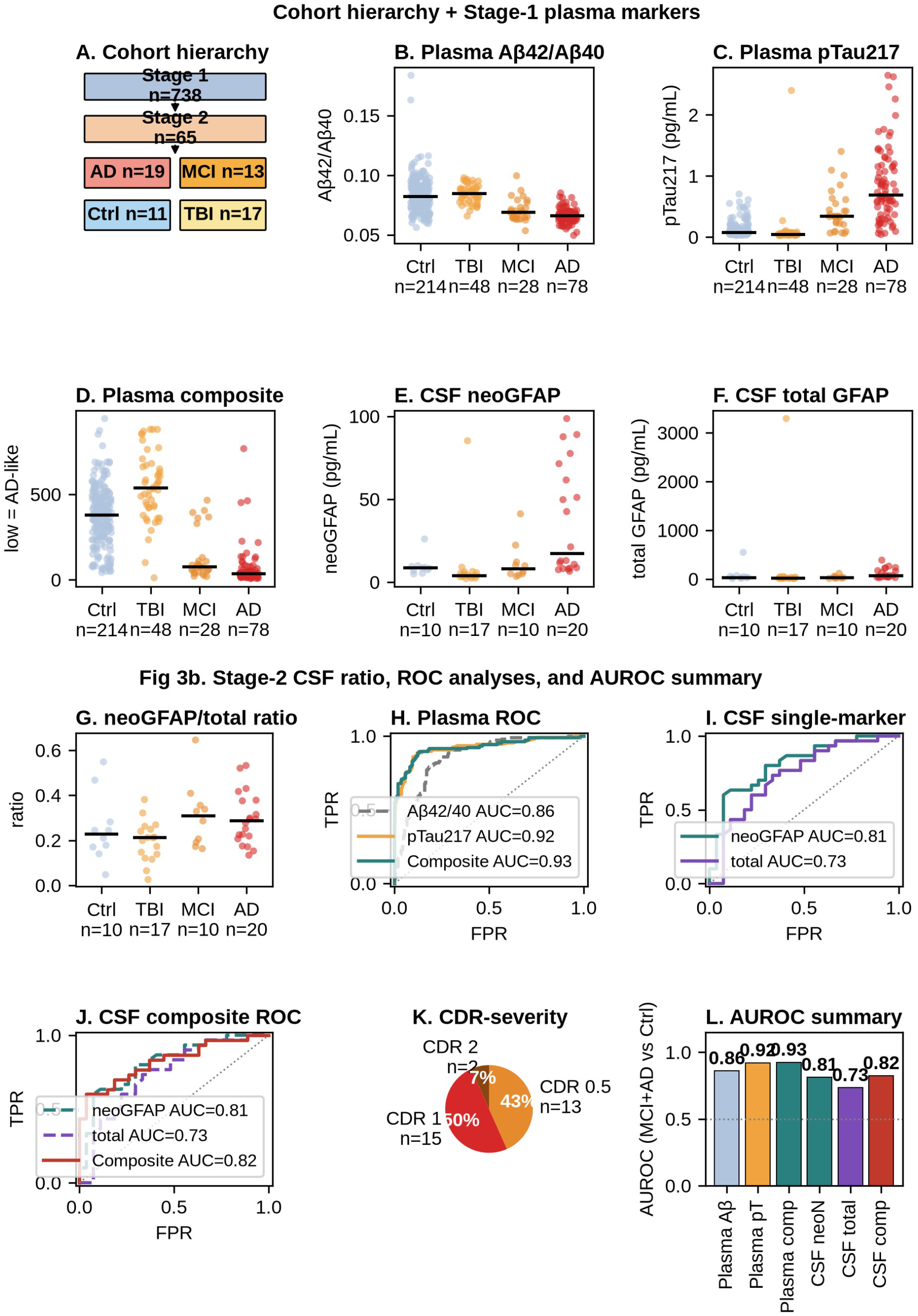
Unified two-stage diagnostic and prognostic biomarker framework, presented in two parts. Figure 3a (top): cohort hierarchy (panel A), Stage-1 plasma gate marker distributions by group (Lumipulse amyloid beta 42/40 and pTau217; panels B–C), the Stage-1 plasma composite (panel D), and Stage-2 CSF single-marker distributions (neoGFAP™, total GFAP; panels E–F). Figure 3b (bottom): Stage-2 CSF proteoform ratio (panel G), Stage-1 plasma ROC (panel H), Stage-2 CSF single-marker and composite ROC (panels I–J), CDR-Global severity mix of Stage-2 subjects (panel K), and AUROC summary bar chart (panel L). Stage-1 gate-positive rates: control 8.*0%, TBI 2.1%, MCI 35.7%, AD 70*.5%.

### Cerebrospinal-fluid head-to-head: neoGFAP™ versus total GFAP

At matched 75th-percentile control-derived cutoffs, CSF neoGFAP™ achieved an AUROC of 0.81 for MCI plus AD versus non-AD (controls plus TBI) compared with 0.73 for total GFAP (Figure 4A, whole cohort n = 57). The paired-bootstrap ΔAUROC was +0.08 (95% CI ™0.00 to +0.16; p = 0.056; McNemar exact p = 0.581): at the whole-cohort level the neoGFAP™ advantage is trend-level and not statistically significant. Within the Stage-2 gate-positive, amyloid-committed subset (Figure 4A′, n = 23), neoGFAP™ dominance became significant—six subjects were correctly classified by neoGFAP™ but misclassified by total GFAP, and none in the reverse direction (McNemar exact p = 0.031; ΔAUROC +0.15). One-sided Mann–Whitney tests showed MCI plus AD greater than non-AD at p = 3×10^−5^ for neoGFAP™ versus p = 1.4×10^−3^ for total GFAP, and Fisher exact gate-enrichment odds ratios were 10.4 for neoGFAP™ versus 4.6 for total GFAP. The signal is thus concentrated in the amyloid-committed subpopulation rather than distributed across the full cohort, and total GFAP provided no classification value that neoGFAP™ did not already capture.

**Figure 4.**
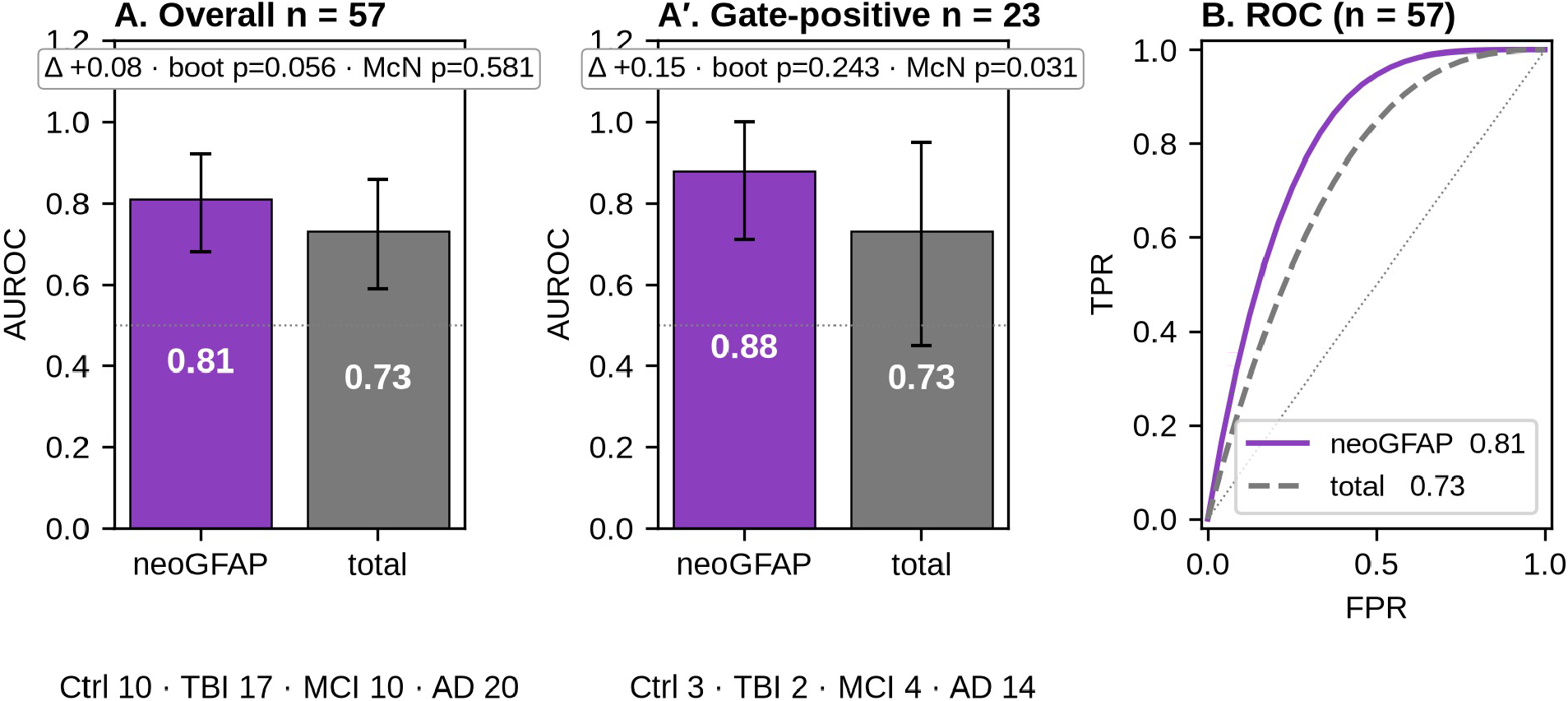
Head-to-head comparison of CSF neoGFAP™ versus CSF total GFAP for the MCI+AD vs non-AD (control + chronic blast TBI) diagnostic task, at two levels of stringency. (A) Overall CSF benchmarking cohort (n = 57): AUROC bars with bootstrap 95% CIs; ΔAUROC = +0.*08, paired-bootstrap p = 0.056, McNemar exact p = 0.581 (trend-level). (A′) Stage-2 gate-positive subset (n = 23): ΔAUROC = +0.15, McNemar exact p = 0.031 (six discordant subjects, all favouring neoGFAP™). (B) Overlaid ROC curves for the overall n = 57 analysis*.

### Diagnostic and prognostic performance across the TBI–MCI–AD continuum

Extending the head-to-head comparison across every clinically meaningful contrast (Supplementary Table S2), three findings emerged. First, for AD versus control, neoGFAP™ (AUROC 0.81) again outperformed total GFAP (0.74; ΔAUROC +0.07), confirming the head-to-head result on a larger contrast. Second, for MCI versus chronic blast-exposed Veterans without cognitive impairment (median CDR-Global 0), neoGFAP™ (0.77) outperformed total GFAP (0.62) by ΔAUROC +0.15. Third, for chronic blast TBI versus control, CSF neoGFAP™ showed paradoxical depletion: TBI subjects had significantly lower CSF neoGFAP™ than controls (AUROC 0.14 in the TBI-greater-than-control direction; Mann–Whitney two-sided p = 0.002), whereas total GFAP showed no significant difference (p = 0.14). Cross-sectionally, CSF neoGFAP™ was elevated 2.16-fold in AD versus control and total GFAP 2.43-fold, both markers were essentially flat in MCI, and in chronic blast TBI neoGFAP™ was depleted to 0.43-fold of control with total GFAP directionally consistent but less extreme (0.48-fold). In the head-to-head AD versus MCI contrast, however, total GFAP outperformed neoGFAP™ (AUROC 0.86 versus 0.79; ΔAUROC ™0.07), the sole primary contrast in which the total-protein assay exceeded the proteoform assay; this is consistent with the graded, class-level information total GFAP provides once AD-versus-non-AD is already resolved by the Stage-1 plasma gate. Four of the five primary diagnostic contrasts survived Benjamini–Hochberg FDR correction (all except the null MCI-diagnostic contrast).

### Chronic-blast paradoxical depletion

The paradoxical CSF depletion of neoGFAP™ in chronic blast TBI (AUROC 0.14; p = 0.002) is the proteoform signature that total GFAP cannot fully resolve. It is consistent with brain-tissue sequestration of aggregated proteoform fragments—analogous to the well-documented CSF amyloid beta 42 drop in AD that reflects parenchymal deposition—and identifies chronic blast TBI as a mechanistically distinct node upstream of the MCI-to-AD trajectory rather than a null result.

### Unbiased Olink proteomic profiling

Genome-wide CSF Olink profiling (2,908 assays) surfaced GFAP as a top-ranked astrocytic hit across disease groups and independent of control-reference choice (young, mid, old, pooled, or the inverted TBI reference). The leading hits across compartments were GFAP (astrocytic), NEFL (neuronal/axonal), MAPT (neuronal-tau), PPIF (mitochondrial), and TREM2 (microglial) (Supplementary Table S3). The five-marker profile revealed a coordinated MCI-plus-AD greater-than-TBI greater-than-control pattern across astrocytic, neuronal, mitochondrial, and microglial compartments, with chronic blast TBI showing attenuated NPX consistent with the CSF neoGFAP™ depletion and a debris-clearance/sequestration model.

### Exploratory subject-level reclassification

In an exploratory reclassification (Supplementary Section S1; leave-one-out cross-validated multinomial logistic regression), a sizeable fraction of clinically diagnosed MCI/AD subjects were biomarker-negative across plasma classifiers, identifying candidates for alternative-etiology workup. Overall classification accuracy rose from 71.1 percent using plasma alone (Set A) to 75.5 percent with added CSF Lumipulse amyloid and tau markers (Set C) and to 79.5 percent with the further addition of age (Set F; n = 380 matched plasma-plus-CSF sample pairs (multiple visits per subject)). Bootstrap 95 percent confidence intervals for the one-versus-rest AUROCs in the fullest model (Set F) were: AD 0.95 (0.92–0.98), control 0.91 (0.88–0.93), MCI 0.79 (0.70– 0.87), and TBI 0.94 (0.91–0.97), with the largest gains for the non-AD classes—consistent with plasma alone carrying a binary AD-burden signal while CSF adds graded class-level information.

On the ProQuantum CSF panel (n = 57), CSF neoGFAP™ achieved an AUROC of 0.76 (95% CI 0.62–0.88) for MCI plus AD versus non-AD, compared with 0.72 (95% CI 0.59–0.86) for total GFAP (ΔAUROC = +0.033, 95% CI ™0.01 to +0.08), preserving the direction and rank ordering of the primary result (per-subject Spearman ρ = 0.84 across platforms). On the same 57-subject cohort, plasma concentrations tracked CSF concentrations for both astroglial markers (per-subject Spearman ρ = 0.63 for neoGFAP™ and ρ = 0.58 for total GFAP; both p = 2×10^−7^ and 2×10^−6^, respectively), and plasma–CSF coupling was strongest within the AD-continuum groups (control, MCI, and AD Spearman ρ = 0.43–0.72 for neoGFAP™), consistent with the two-stage plasma-to-CSF diagnostic scheme. In head-to-head classification of MCI plus AD versus Control plus TBI, plasma neoGFAP™ (with or without plasma total GFAP) was directionally equivalent to plasma pTau217 (AUROCs 0.90 versus 0.92; paired ΔAUROC = ™0.02, 95% CI ™0.13 to +0.09), and outperformed plasma Aβ42/40 by 6–7 AUROC points as a solo classifier (AUROCs 0.90 versus 0.84; paired ΔAUROC = +0.06, 95% CI ™0.07 to +0.19), positioning plasma neoGFAP™ as a candidate astroglial-proteoform readout alongside the plasma amyloid/tau reference markers in this population.

## Discussion

This study delivers three convergent lines of evidence that the calpain-cleaved neoGFAP™ proteoform—not total GFAP—may be the diagnostically informative astroglial biomarker across the healthy-control (HC)/TBI– MCI–AD continuum. First, within a formal two-level plasma-to-CSF gating scheme, neoGFAP™ outperformed total GFAP for AD confirmation on multiple statistical tests, including a paired McNemar comparison that ruled out any complementary value of total GFAP (Figure 4) (six versus zero discordant pairs; p = 0.031 in the gate-positive subset). Second, extended across every clinically meaningful contrast, neoGFAP™ was stronger for MCI versus chronic blast-exposed Veterans without cognitive impairment (ΔAUROC +0.15). Third, unbiased Olink proteomics across astrocytic, neuronal, mitochondrial, and microglial compartments independently confirmed the coordinated MCI-plus-AD greater-than-TBI greater-than-control pattern.

The split whole-cohort versus gate-positive result is central to interpretation. At the whole-cohort level (n = 57) the neoGFAP™ advantage over total GFAP is trend-level (ΔAUROC +0.08; McNemar p = 0.581); it becomes McNemar-significant only within the Stage-1 amyloid-positive subset (n = 23; six discordant pairs, all favouring neoGFAP™; p = 0.031). The proteoform-specific diagnostic signal therefore emerges specifically in the amyloid-committed subpopulation, exactly where a graded severity readout is clinically useful, and is consistent with the two-stage design in which Stage 2 is applied only after Stage-1 amyloid triage.

The paradoxical CSF depletion of neoGFAP™ in chronic blast TBI merits particular discussion. Rather than a null result, this depletion tracks the long-standing observation in AD that CSF amyloid beta 42 drops precisely because it is sequestered in parenchymal plaques—the biomarker signal moves from fluid to tissue. Calpaingenerated neoGFAP™ fragments are known to form filamentous aggregates, and Alexander disease demonstrates that GFAP proteoforms readily template Rosenthal fibers.^2^ If neoGFAP™ similarly aggregates in chronic blast TBI tissue, CSF depletion becomes a mechanistically coherent biomarker of chronic blast TBI-to-AD transition rather than an absence of pathology.

### Interpretation: an astroglial-proteoform axis for A/T/N

Our results operationalize the central insight that an AD-like pattern of tau accumulation as a function of amyloid beta is observed only in individuals with high GFAP—astroglial reactivity is the necessary gating event, not merely a correlated biomarker.^6,7,8,9,15^ All of these studies were performed with assays targeting total GFAP, which aggregate intact GFAP with multiple proteoforms.^10^ Our data suggest that the biologically active discriminant signal resides specifically in the calpain-cleaved neoGFAP™ proteoform, and that proteoform-resolved assays should therefore sharpen operationalization of this framework for clinical use. Accordingly, with replication in a larger dataset, a proteoform-resolved astroglial axis (G) may be considered for addition to the A/T/N framework^5^ as A/T/N/G, particularly for TBI-exposed populations. Because Veterans with TBI carry an approximately two- to four-fold elevated risk of dementia,^19^ a biomarker that separates MCI from chronic blast exposure without cognitive impairment (ΔAUROC +0.15) supports further evaluation for Department of Defense and Veterans Affairs surveillance programs.

### Comparison with prior work

Our CSF AD-reference panel was measured on the Fujirebio Lumipulse G platform—the same platform underlying the FDA-cleared Lumipulse amyloid beta 42/40 CSF assay—and our CSF benchmarking sits within the published Lumipulse-CSF performance envelope; the May 2025 FDA marketing clearance of the Lumipulse plasma pTau217/amyloid beta 42 ratio established the first FDA-cleared blood test for AD diagnosis.^16^ For the Stage-1 plasma gate, published Lumipulse plasma amyloid beta 42/40 AUROCs for abnormal amyloid-PET cluster around 0.78–0.82,^12,13^ and our Stage-1 ratio (AUROC 0.93) sits at the upper end of this range. This is consistent with the Lumipulse plasma test’s strong performance in unselected populations, but as Rosen-Lang and colleagues showed, that performance does not carry over to Veterans with prior TBI,^11^ which is precisely the population the Stage-2 CSF neoGFAP™ step is intended to recover. For the chronic blast TBI arm, the closest published comparators are the Peskind/Li studies in this Veteran population: cerebrocerebellar glucose hypometabolism on FDG-PET after repetitive blast exposure,^1^ and declining CSF amyloid beta in middle-aged Veterans with blast mild TBI while CSF pTau181/tTau remained relatively constant,^14^ with the interpretive caveats of that CSF literature set out in an accompanying editorial commentary;^17^ our CSF benchmarking reproduces the CSF features and adds a proteoform-resolved astroglial axis, with neoGFAP™ discriminating chronic blast TBI from controls more strongly than total GFAP. Recent plasma-astroglial reports converge on GFAP as a central diagnostic and prognostic axis: higher baseline serum GFAP and neurofilament light were associated with a greater rate of brain atrophy over five years after TBI,^18^ plasma GFAP is elevated approximately a decade before expected symptom onset in autosomal dominant AD,^20^ and plasma GFAP mediated the effect of amyloid beta on tau burden;^8^ our neoGFAP™ results extend this literature by isolating the calpain-cleavage proteoform, the most TBI-relevant fragment.

### Limitations

Several limitations should remain explicit. The CSF benchmarking cohort is modest (n = 57), particularly within the TBI (n = 17) and MCI (n = 10) strata, and the whole-cohort neoGFAP™ advantage over total GFAP is only trend-level; the McNemar-significant result derives from the smaller gate-positive subset (n = 23) and requires prospective replication. Some Olink NPX analyses were sample-level rather than subject-level.

### Cohort composition and demographic confounding

The cohort reflects the source populations rather than a matched case-control design: the TBI arm is drawn from Department of Defense and Veterans Affairs deployed-Veteran registries (younger, predominantly male) while the MCI and AD arms are drawn from community memory-disorder cohorts (older, more sex-balanced) (Table 1b). Age and sex are therefore almost perfectly confounded with diagnostic category, and the diagnostic AUROCs reported here cannot be cleanly attributed to disease biology independent of demographic effects. A sensitivity analysis restricted to age- and sex-overlapping subjects would be underpowered; a properly powered replication requires a prospective matched design recruiting age- and sex-balanced arms across TBI, MCI, AD, and healthy controls. Within-arm longitudinal analyses, where each subject serves as their own control, are less affected. TBI-exposure history was not ascertained in the MCI and AD arms, and the chronic blast TBI arm was cognitively unimpaired (median CDR-Global 0), so the MCI-versus-TBI contrast reflects discrimination of cognitive impairment in a memory-disorder cohort from chronic blast exposure without cognitive impairment rather than discrimination of MCI with a TBI history from MCI without one. Because this is a secondary analysis of archived data with cross-cohort composition differences, findings should be interpreted as hypothesis-generating rather than definitive for clinical deployment.

The results support further prospective evaluation of CSF neoGFAP™-based staging in a matched-cohort design across the TBI–MCI–AD continuum.

### Future work

The immediate next experiment is to run the same neoGFAP™ and total GFAP immunoassays on the banked plasma aliquots already matched to this cohort, enabling a plasma-only multi-class reclassifier and a direct head-to-head against the FDA-cleared Lumipulse plasma pTau217/amyloid beta 42 ratio; longitudinal time-to-event analyses and extension to capillary sampling formats are reserved for prospective cohorts.

### Conclusion

Across a two-stage plasma-to-CSF scheme in Veterans, the calpain-cleaved neoGFAP™ proteoform outperformed total GFAP for diagnostic and prognostic classification along the TBI–MCI–AD continuum, most clearly within the amyloid-committed subpopulation and for the MCI-versus-TBI differential. These findings support adding an astroglial-proteoform axis to A/T/N biomarker frameworks in high-risk populations, pending prospective, demographically matched validation.

## Declarations

### Ethical Oversight

Archived CSF and plasma samples were obtained under Institutional Review Board approval at the VA Puget Sound Health Care System (ADTBI IRB #1653649) from the VA Northwest Mental Illness Research, Education, and Clinical Center (VA NW MIRECC) Sample and Data Biorepository. That Institutional Review Board oversight covered the entire study, including all cohorts and all human samples and human data reported here.

### Informed Consent

All participants (or their legally authorized representatives) provided written informed consent for biorepository collection and future research use under the governing Institutional Review Board protocol (ADTBI IRB #1653649).

## Data availability

De-identified data from this study are available upon reasonable request to the corresponding author, subject to VA NW MIRECC biorepository governance and ADTBI IRB #1653649 restrictions.

## Acknowledgements

The authors thank the participants and families whose participation made this study possible. We thank the VA Northwest MIRECC, VA Puget Sound Health Care System, and University of Washington School of Medicine clinical research teams for cohort development, sample collection, and clinical annotation. AI-assistance disclosure: The authors used Perplexity Computer (Perplexity AI, San Francisco, CA) as an AI research assistant for literature synthesis, tracked-change reconciliation across coauthor revisions, figure drafting, and manuscript-preparation support during drafting of this manuscript. All scientific content, biomarker data analyses, interpretations, conclusions, and final wording were reviewed, verified, and approved by the human authors, who take full responsibility for the integrity and accuracy of the work.

## Authors’ Contributions

KKW, RG, AP, JBW, ABW, EP, and WEH conceptualized the work, KKW, WEH, SB, and MM developed the methodology, SB, MM, GC, KB, EE, DJ, and KT led the analysis of biomarkers in the deidentified samples, MK, SB, MM, and SJ performed the ProQuantum immunoassays, GGL and EP contributed to cohort development, sample collection, and clinical annotation, TK and FO provided therapeutic-development context, KR provided project administration and regulatory coordination, WEH performed the formal data analysis, and WEH led manuscript writing with assistance from RG, AP, JBW, ABW, EP, KKW, SJ, RB, AD, CP, and GM.

## Clinical Trial Registration

This was a cross-sectional, retrospective, single-site observational biomarker study; no intervention was prospectively assigned to participants. Clinical trial registration therefore does not apply.

## References

1. Peskind ER, Petrie EC, Cross DJ, et al. Cerebrocerebellar hypometabolism associated with repetitive blast exposure mild traumatic brain injury in 12 Iraq war Veterans with persistent post-concussive symptoms. Neuroimage. 2011;54(Suppl 1):S76–S82. doi:10.1016/j.neuroimage.2010.04.008

2. Yang Z, Arja RD, Zhu T, et al. Characterization of calpain and caspase-6-generated glial fibrillary acidic protein breakdown products following traumatic brain injury and astroglial cell injury. Int J Mol Sci. 2022;23(16):8960. doi:10.3390/ijms23168960

3. Robertson CS, Salinas Martinez F, McQuillan LE, Williamson J, Lamb DG, Wang KKW, et al. Serial measurements of serum glial fibrillary acidic protein in moderate-severe traumatic brain injury: potential utility in providing insights into secondary insults and long-term outcome. J Neurotrauma. 2024;41(1-2):73–90. doi:10.1089/neu.2023.0111

4. Wang KKW, Barton DJ, McQuillan LE, et al. Parallel cerebrospinal fluid and serum temporal profile assessment of axonal injury biomarkers neurofilament-light chain and phosphorylated neurofilament-heavy chain: associations with patient outcome in moderate-severe traumatic brain injury. J Neurotrauma. 2024;41(13-14):1609–1627. doi:10.1089/neu.2023.0449

5. Jack CR Jr, Bennett DA, Blennow K, et al. NIA-AA Research Framework: toward a biological definition of Alzheimer’s disease. Alzheimers Dement. 2018;14(4):535–562. doi:10.1016/j.jalz.2018.02.018

6. Bellaver B, Povala G, Ferreira PCL, et al. Astrocyte reactivity influences amyloid-β effects on tau pathology in preclinical Alzheimer’s disease. Nat Med. 2023;29(7):1775–1781. doi:10.1038/s41591-023-02380-x

7. Ferrari-Souza JP, Ferreira PCL, Bellaver B, et al. Astrocyte biomarker signatures of amyloid-β and tau pathologies in Alzheimer’s disease. Mol Psychiatry. 2022;27(11):4781–4789. doi:10.1038/s41380-022-01716-2

8. Pereira JB, Janelidze S, Smith R, et al. Plasma GFAP is an early marker of amyloid-β but not tau pathology in Alzheimer’s disease. Brain. 2021;144(11):3505–3516. doi:10.1093/brain/awab223

9. Chatterjee P, Pedrini S, Stoops E, et al. Plasma glial fibrillary acidic protein is elevated in cognitively normal older adults at risk of Alzheimer’s disease. Transl Psychiatry. 2021;11(1):27. doi:10.1038/s41398-020-01137-1

10. Gogishvili D, Honey MIJ, Verberk IMW, et al. The GFAP proteoform puzzle: how to advance GFAP as a fluid biomarker in neurological diseases. J Neurochem. 2025;169(1):e16226. doi:10.1111/jnc.16226

11. Rosen-Lang Y, Vrillon A, Pasternak S, et al; Department of Defense Alzheimer’s Disease Neuroimaging Initiative (DOD ADNI) Investigators and the Department of Defense Alzheimer’s Disease Blood-Testing Initiative (DOD ADBI). Prior traumatic brain injury and Alzheimer disease blood biomarkers. JAMA Neurol. 2026;83(8):759–768. doi:10.1001/jamaneurol.2026.2042

12. Schindler SE, Petersen KK, Bollack A, et al. Head-to-head comparison of leading blood tests for Alzheimer’s disease pathology. Alzheimers Dement. 2024;20(11):8074–8096. doi:10.1002/alz.14315

13. Pilotto A, Quaresima V, Trasciatti C, et al. Plasma p-tau217 in Alzheimer’s disease: Lumipulse and ALZpath SIMOA head-to-head comparison. Brain. 2025;148(2):408–415. doi:10.1093/brain/awae368

14. Li G, Iliff J, Shofer J, Mayer CL, Meabon J, Cook D, et al. CSF β-Amyloid and Tau Biomarker Changes in Veterans With Mild Traumatic Brain Injury. Neurology. 2024;102(7):e209197. doi:10.1212/WNL.0000000000209197

15. Benedet AL, Milà-Alomà M, Vrillon A, et al. Differences between plasma and cerebrospinal fluid glial fibrillary acidic protein levels across the Alzheimer disease continuum. JAMA Neurol. 2021;78(12):1471–1483. doi:10.1001/jamaneurol.2021.3671

16. US Food and Drug Administration. FDA clears first blood test used in diagnosing Alzheimer’s disease. FDA News Release. May 16, 2025. Accessed July 2026. https://www.fda.gov/news-events/press-announcements/fda-clears-first-blood-test-used-diagnosing-alzheimers-disease

17. Alosco ML, Tartaglia MC. Repetitive Blast Injury and CSF Alzheimer Disease Biomarkers: Navigating a Complex Narrative. Neurology. 2024;102(7):e209294. doi:10.1212/WNL.0000000000209294

18. Shahim P, Pham DL, van der Merwe AJ, et al. Serum NfL and GFAP as biomarkers of progressive neurodegeneration in traumatic brain injury. Alzheimers Dement. 2024;20(7):4663–4676. doi:10.1002/alz.13898

19. Barnes DE, Byers AL, Gardner RC, et al. Association of mild traumatic brain injury with and without loss of consciousness with dementia in US military veterans. JAMA Neurol. 2018;75(9):1055–1061. doi:10.1001/jamaneurol.2018.0815

20. Chatterjee P, Vermunt L, Gordon BA, et al. Plasma glial fibrillary acidic protein in autosomal dominant Alzheimer’s disease: associations with Aβ-PET, neurodegeneration, and cognition. Alzheimers Dement. 2023;19(7):2790–2804. doi:10.1002/alz.12879

