## Supplemental for "Proteoform-resolved neoGFAP^™^ as a diagnostic and prognostic biomarker across the TBI–MCI–AD continuum in Veterans"

**Frame reconciliation.** The three cohort frames used in this manuscript are non-nested and have distinct denominators:

**Frame 1 — Stage-1 plasma screening cohort** (Table 1, n = 367 unique subjects): Control n = 213; MCI n = 28; AD n = 78; chronic blast TBI (TBI-T + TBI-C + TBI-N) n = 48. Sum: 213 + 28 + 78 + 48 = 367.

**Frame 2 — CSF Stage-2 panel-complete benchmarking cohort** (Table 1b, n = 57 unique subjects): Control n = 10; chronic blast TBI n = 17; MCI n = 10; AD n = 20. Every subject has paired CSF neoGFAP™ AND CSF total GFAP AND plasma pTau217 measurements.

**Frame 3 — Reclassification analysis frame** (Supplementary Section S1, n = 380 matched plasma + CSF pairs): This is a per-visit, not per-subject, count. The 380 pairs derive from a subset of Stage-1 subjects who have both plasma AND CSF measured, allowing multiple visits per subject where longitudinal CSF sampling occurred. Approximately 220 unique subjects contribute the 380 pairs (median ~1.7 visits per subject; range 1–4).

The apparent discrepancy between Frame 1 MCI n = 28 (unique subjects, Stage-1) and Frame 2 MCI n = 10 (panel-complete CSF) reflects the panel-completeness gate: only 10 of the 28 unique-subject MCI cases had both a CSF neoGFAP™ AND a CSF total GFAP measurement available at Stage-2 timing. Similar attrition applies to Control (213 → 10), AD (78 → 20), and TBI (48 → 17) arms, and reflects the natural falloff between plasma screening (widely available) and CSF sampling (available only where clinically indicated).

#### Supplementary Figures

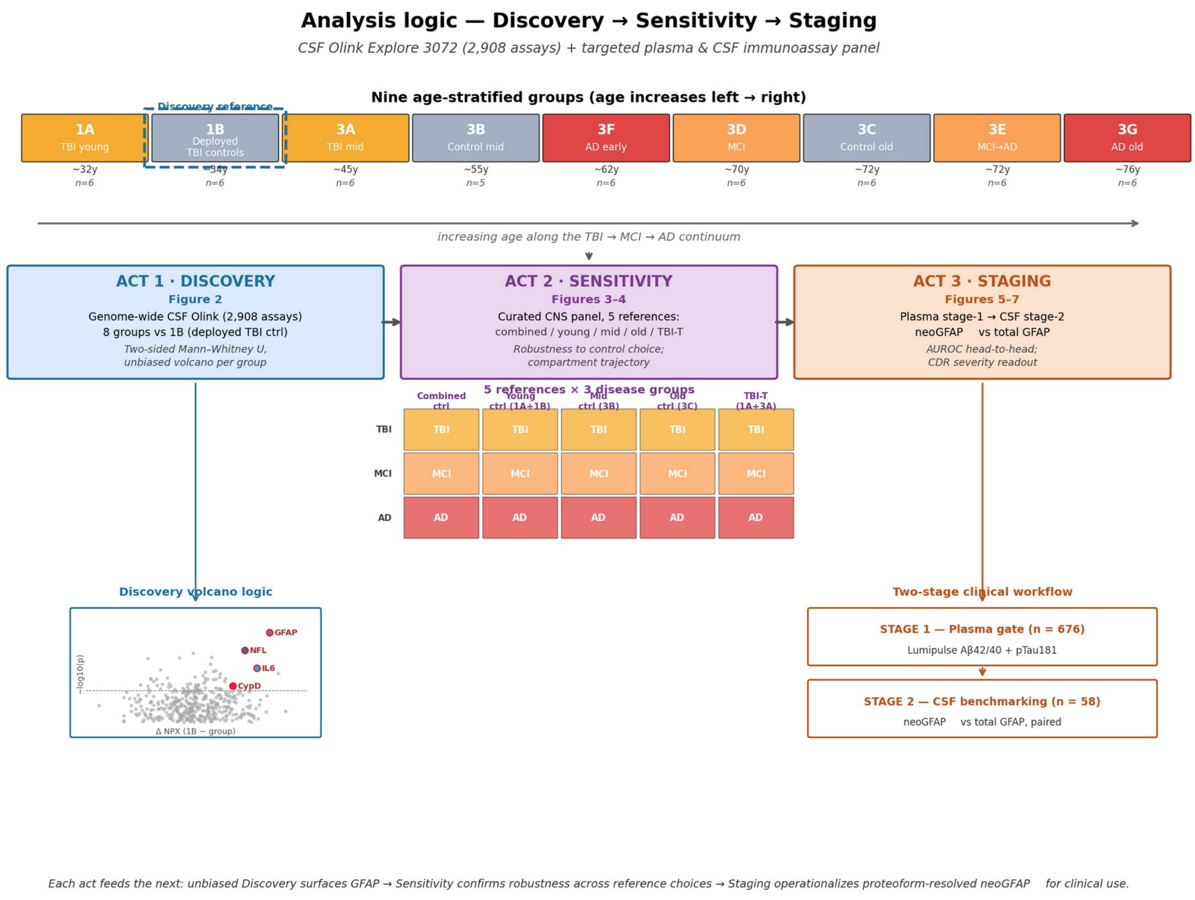


*Supplementary Figure S1. Analysis logic, study workflow, and biomarker downselection. The study spans nine age-stratified subject groups along the chronic-blast-TBI → MCI → AD continuum, organized in three acts (Discovery, Sensitivity, Staging) with explicit data-driven downselection at each act.*

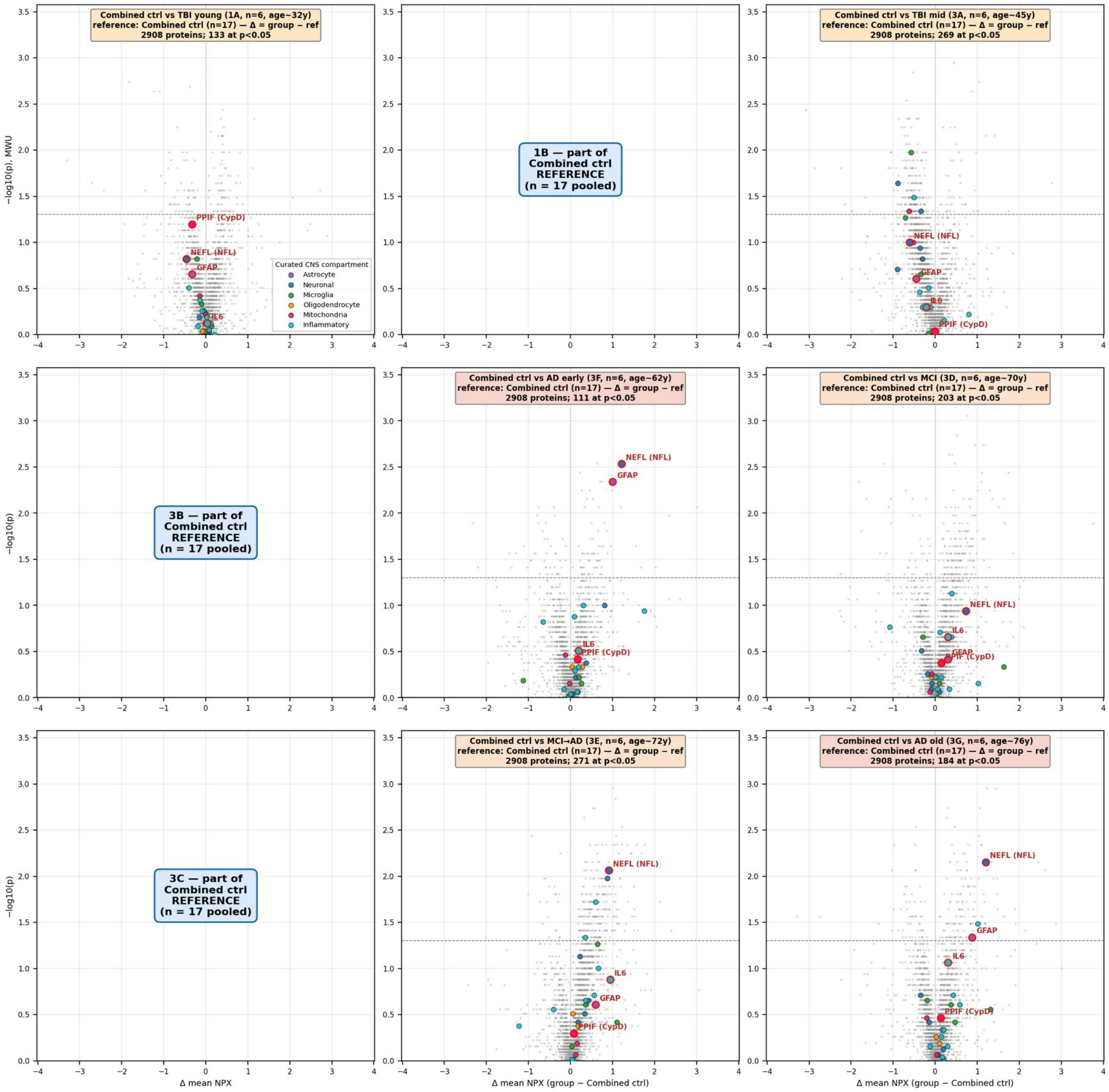


*Supplementary Figure S2. Discovery — genome-wide CSF Olink Explore 3072 volcano plots (all 2,908 assays passing QC), including the sensitivity-to-control-choice analysis using combined controls as the reference. GFAP, NEFL, IL6, and PPIF remain on the elevated side across disease groups regardless of control-reference choice.*

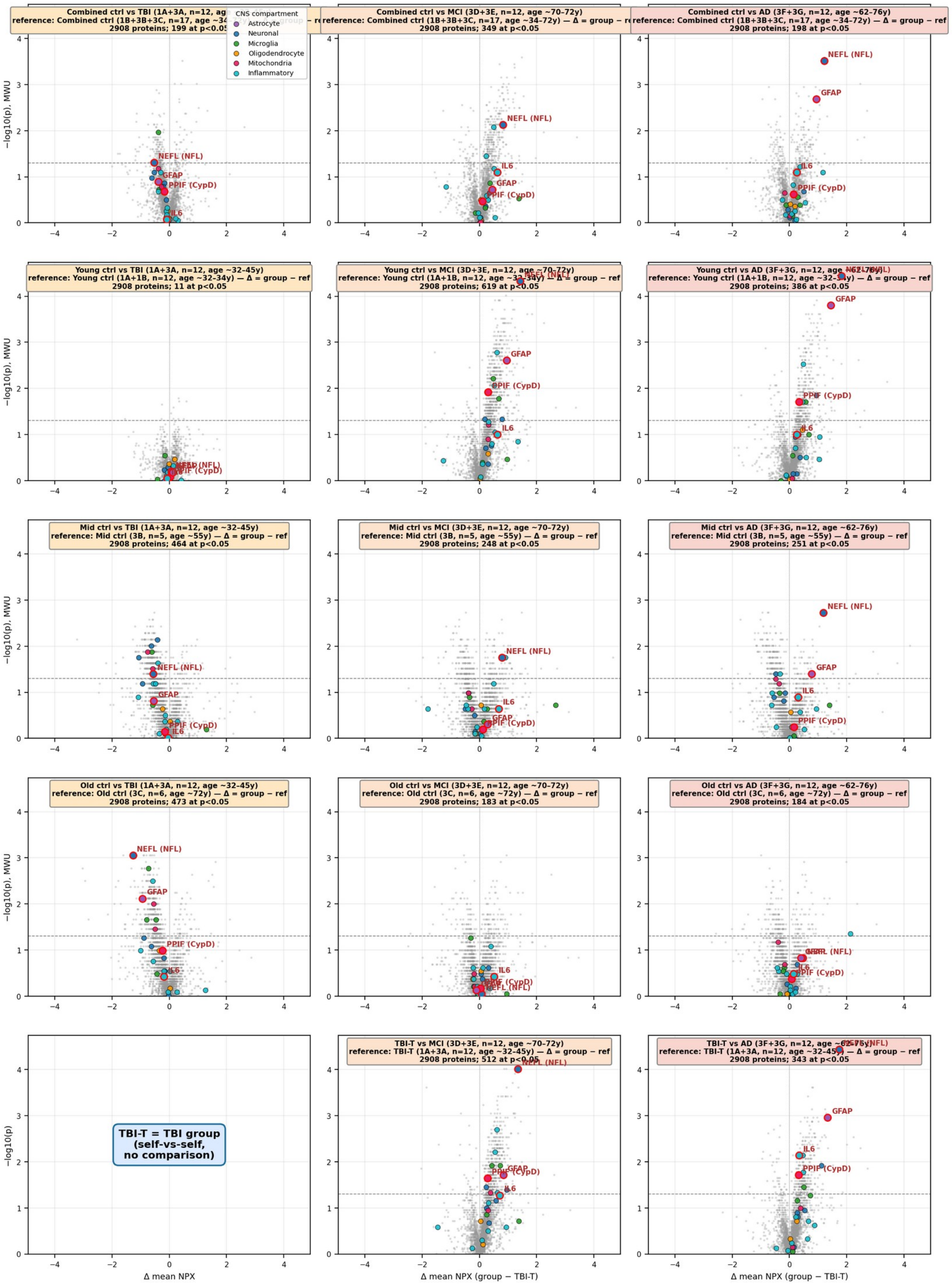

*Supplementary Figure S3. Sensitivity — genome-wide CSF Olink Explore 3072 grid across five reference choices (combined, young, mid, old controls, and the inverted TBI reference) by disease group (TBI, MCI, AD). GFAP and NEFL remain elevated in MCI and AD across all reference choices.*

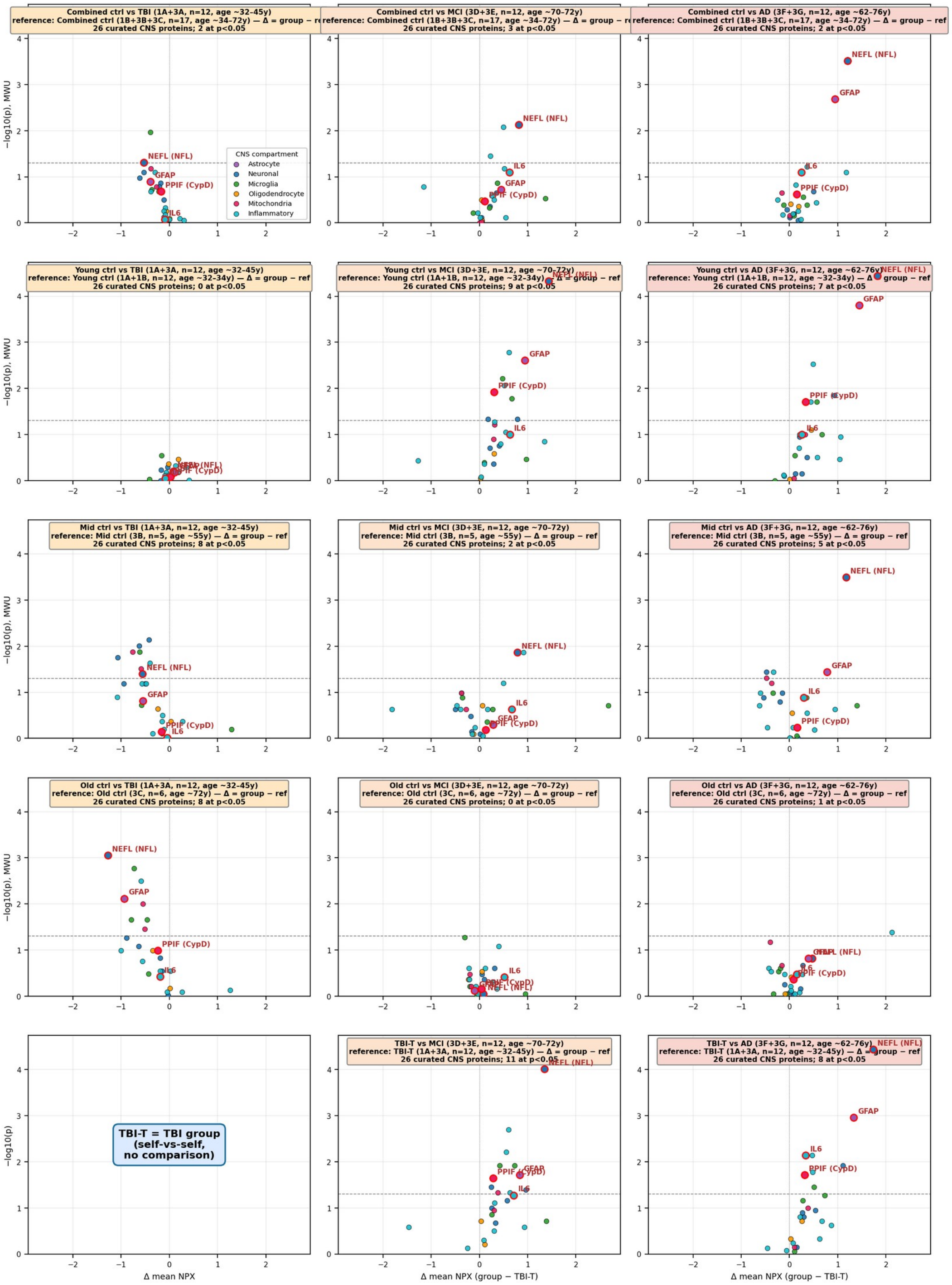


*Supplementary Figure S4. Compartment-stratified summary across every Discovery and Sensitivity comparison. Top: heatmap of significant-protein counts per CNS compartment for each reference-by-disease pair. Bottom: GFAP-specific delta NPX across all comparison columns; GFAP is consistently elevated in MCI and AD across every reference choice.*

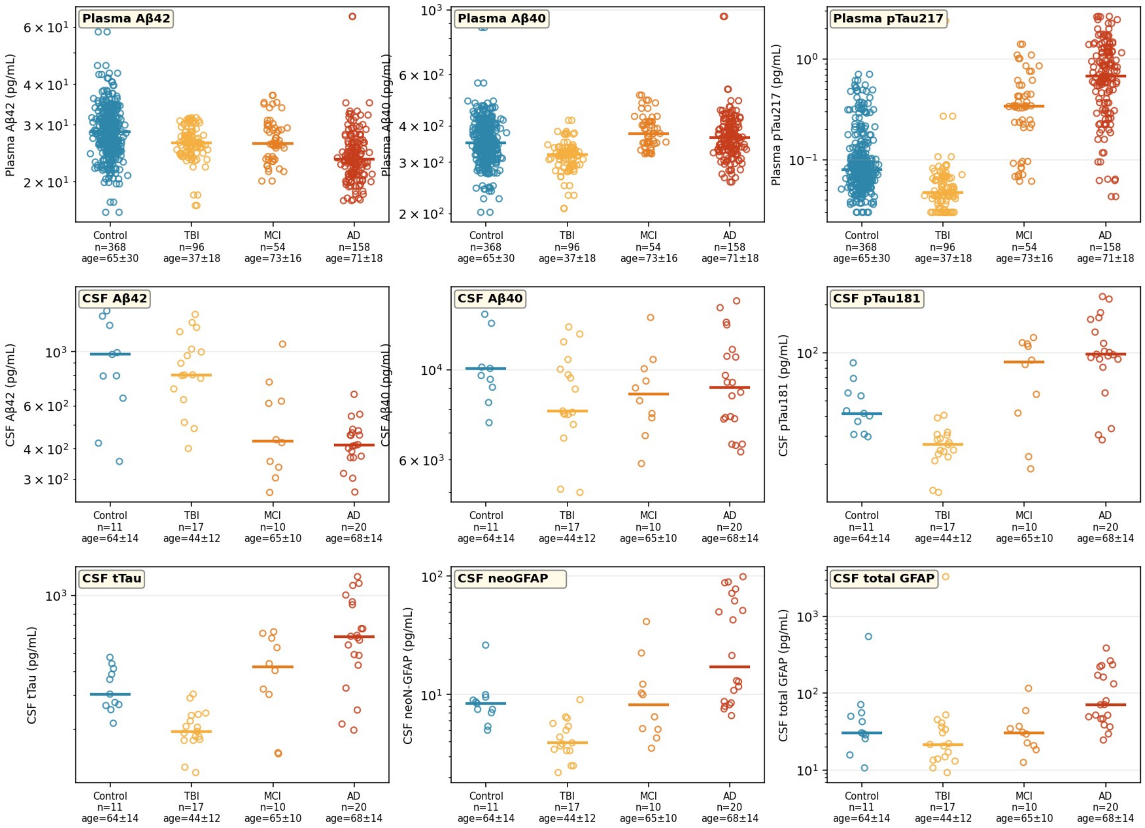


*Supplementary Figure S5. Raw biomarker values by group across all nine assays used in this study (three plasma + six CSF). Open circles represent individual subjects; horizontal bars indicate group medians (log scale). Plasma and Lumipulse CSF assays are Fujirebio Lumipulse G; CSF neoGFAP™ and total GFAP are proprietary Gryphon MSD electrochemiluminescence proteoform assays.*

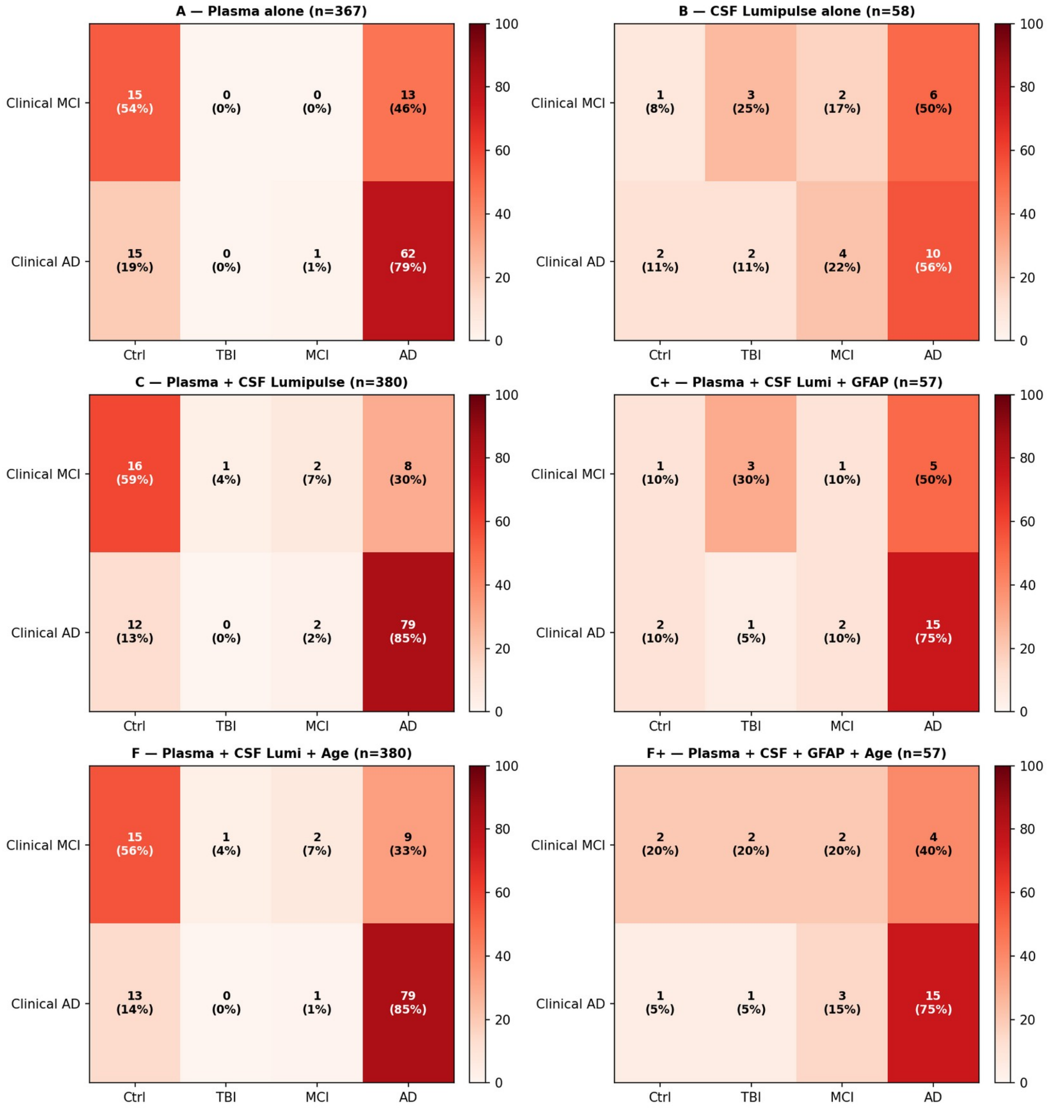


*Supplementary Figure S6. Six-panel heatmap of clinically diagnosed MCI/AD subjects redistributed across the four predicted classes under leave-one-out cross-validation, for the reclassification feature sets described in Supplementary Section S1.*

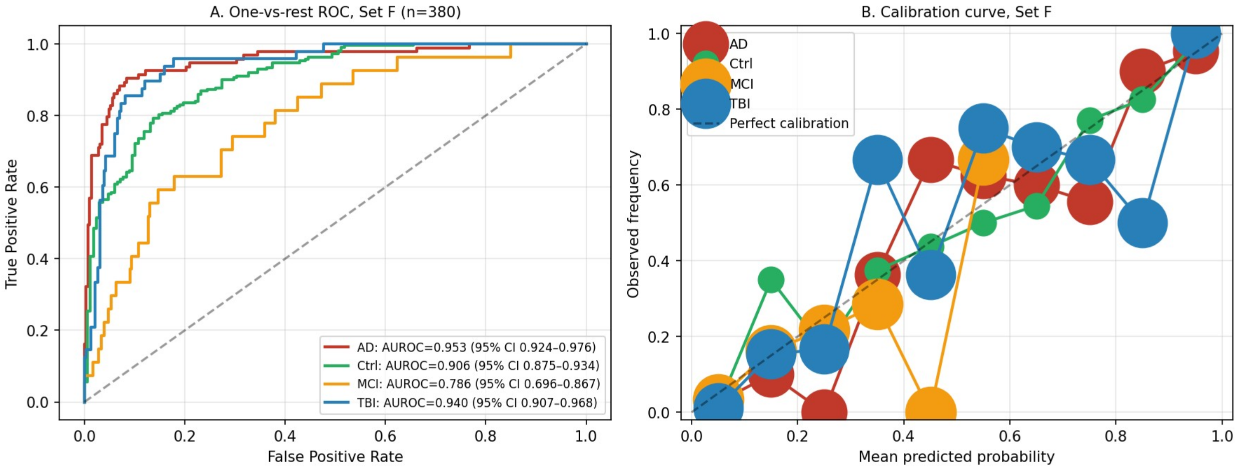


*Supplementary Figure S7. One-versus-rest ROC curves for Set F (plasma + CSF + age, n = 380) under leave-one-out cross-validation, with AUROC point estimates and 2,000-replicate bootstrap 95% confidence intervals.*

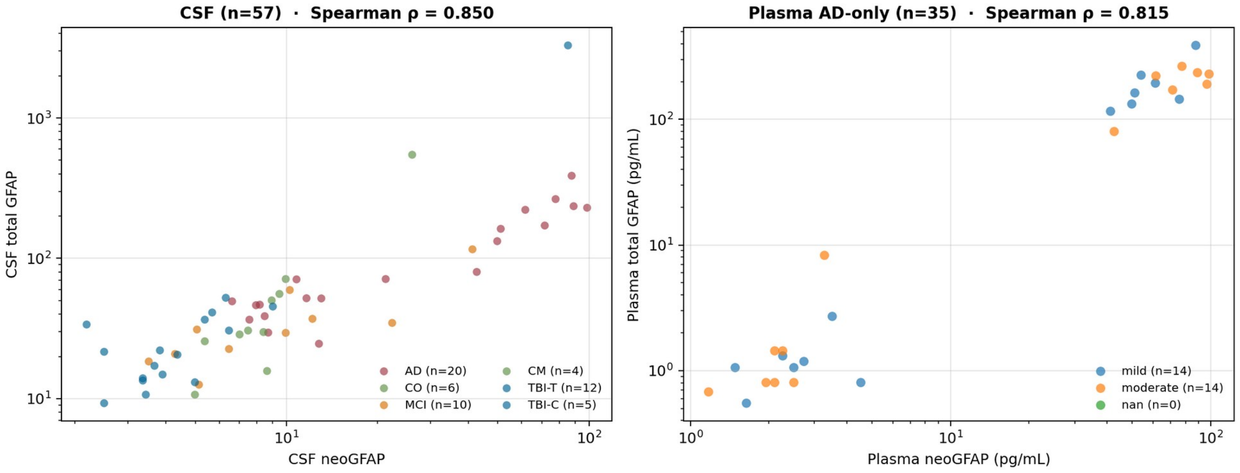


*Supplementary Figure S8. neoGFAP™ versus total GFAP scatter on log10 axes. Left: CSF (n = 57) colored by diagnostic group (Spearman correlation 0.850 overall). Right: plasma AD-only subset (n = 35).*

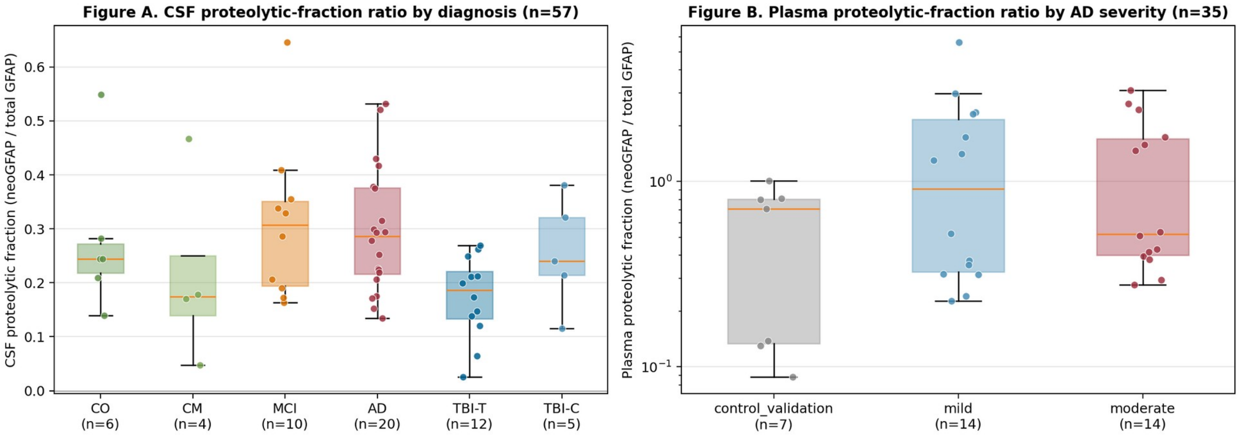


*Supplementary Figure S9. Proteolytic-fraction ratio (neoGFAP™ / total GFAP) by group. CSF (left, n = 57) shows the MCI/AD-high versus TBI-low pattern; plasma AD-only (right, n = 35) is shown for reference.*

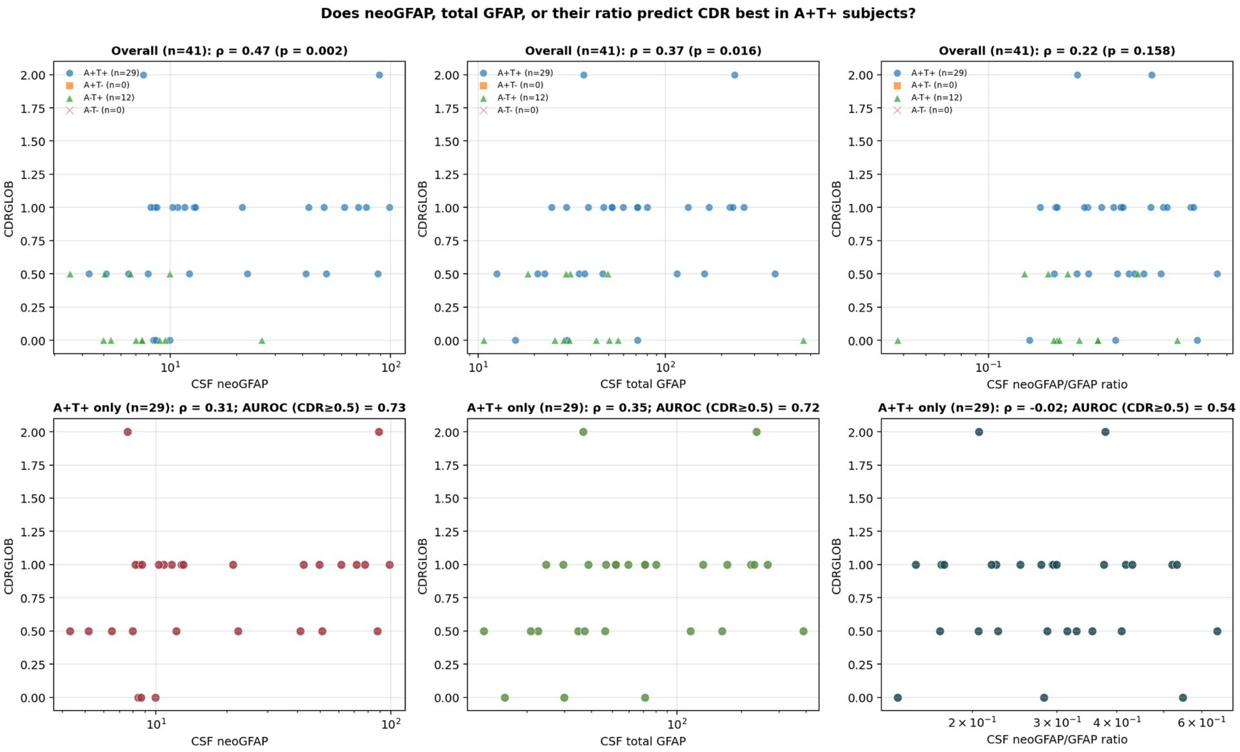


*Supplementary Figure S10. CDR-Global versus each of three CSF features (neoGFAP™, total GFAP, and their ratio) across the cohort, stratified by amyloid/tau status.*

#### Supplementary Tables

***Supplementary Table S1.*** *Stage-1 plasma-gate assignment by clinical group (n = 738). Cutoffs re-derived from the Lumipulse plasma control pool. Stage 0 = neither marker abnormal; Stage 1 = one marker abnormal (borderline); Stage 2 = both markers abnormal at the low cutoffs; Stage 3 = pTau217 above the high cutoff with low amyloid beta 42.*

| Group | Stage 0 | Stage 1 | Stage 2 | Stage 3 | Total | Gate-positive (Stage ≥ 2) |
|---|---|---|---|---|---|---|
| Ctrl | 244 | 152 | 30 | 4 | 430 | 7.9% |
| TBI | 54 | 40 | 2 | 0 | 96 | 2.1% |
| MCI | 12 | 24 | 8 | 12 | 56 | 35.7% |
| AD | 6 | 40 | 14 | 96 | 156 | 70.5% |
| All | 316 | 256 | 54 | 112 | 738 | — |

***Supplementary Table S2.*** *Diagnostic and prognostic AUROC across the TBI–MCI–AD continuum. Mann–Whitney p values are nominal (uncorrected). Under Benjamini–Hochberg FDR, four of five primary contrasts survive at alpha = 0.05 (all except the null MCI-diagnostic contrast).*

| Comparison | n | AUROC neoGFAP™ | AUROC total GFAP | ΔAUROC | MW p (neoGFAP™) | MW p (total) | BH-FDR q | Holm p |
|---|---|---|---|---|---|---|---|---|
| TBI diagnostic | 27 | 0.144 | 0.332 | −0.187 | $1.9\times10^{-3}$ | 0.145 | 0.0095 | 0.0095 |
| MCI diagnostic | 20 | 0.495 | 0.436 | +0.059 | 1.000 | 0.647 | 1.000 | 1.000 |
| AD diagnostic | 30 | 0.814 | 0.743 | +0.070 | $4.7\times10^{-3}$ | 0.029 | 0.0117 | 0.0187 |
| AD vs MCI diagnostic | 30 | 0.790 | 0.855 | −0.065 | 0.011 | $1.9\times10^{-3}$ | 0.019 | 0.0342 |

| | | | | | | | | |
|---|---|---|---|---|---|---|---|---|
| MCI vs TBI diagnostic | 27 | 0.774 | 0.618 | +0.156 | 0.021 | 0.328 | 0.026 | 0.0416 |

***Supplementary Table S3.*** *Top-ranked CNS-compartment Olink markers from the unbiased CSF proteomic screen. Median NPX by group; Kruskal–Wallis p across Control/TBI/MCI+AD; Mann–Whitney p for AD vs Control and AD vs TBI. GFAP is the top-ranked astrocytic marker.*

| Marker | Cell type | Median Ctrl | Median TBI | Median MCI+AD | Kruskal-Wallis p | MW p AD vs Ctrl | MW p AD vs TBI |
|---|---|---|---|---|---|---|---|
| GFAP | Astrocytic | −0.47 | −1.03 | −0.00 | $2.5\times10^{-4}$ | $1.08\times10^{-3}$ | $3.09\times10^{-4}$ |
| NEFL | Neuronal/ axonal | −0.97 | −1.48 | +0.05 | $1.35\times10^{-7}$ | $1.18\times10^{-5}$ | $8.23\times10^{-7}$ |
| MAPT | Neuronal (tau) | −0.80 | −1.10 | +0.07 | $3.37\times10^{-3}$ | $1.04\times10^{-2}$ | $1.50\times10^{-3}$ |
| PPIF | Mitochondrial (CypD) | −0.19 | −0.41 | −0.07 | $2.12\times10^{-2}$ | $1.04\times10^{-1}$ | $2.68\times10^{-3}$ |
| TREM2 | Microglial | −0.27 | −0.70 | +0.04 | $2.23\times10^{-2}$ | $7.03\times10^{-2}$ | $3.23\times10^{-3}$ |

***Supplementary Table S4.*** *Prior literature on GFAP staging — emphasis on GFAP-required-for-cognitive-decline in amyloid-positive individuals.*

| Reference | Cohort | Key finding | Relevance |
|---|---|---|---|
| Bellaver et al., Nat Med 2023 | Cognitively unimpaired Aβ+ | AD-like tau-PET pattern only in high-GFAP individuals | Astroglial reactivity gates Aβ→tau axis |
| Ferrari-Souza et al., Mol Psychiatry 2022 | Preclinical AD | Plasma GFAP mediates Aβ → p-tau181 | Astrogliosis precedes tauopathy |
| Pereira et al., Brain 2021 | A+T−/A+T+ stages | GFAP elevated only in Aβ+; predicts decline | GFAP threshold required for decline |
| Chatterjee et al., Transl Psychiatry 2021 | Cognitively unimpaired elderly | Plasma GFAP separates Aβ+ up to 10 yr pre-dx | Early staging utility |
| Benedet et al., JAMA Neurol 2021 | ADNI | Plasma GFAP tracks Aβ independent of p-tau | Astroglial axis of A/T/N(/G) |
| Gogishvili et al., J Neurochem 2025 | Multi-platform | Total GFAP conflates proteoforms | Rationale for proteoform-resolved assays |
| Shahim et al., Alzheimers Dement 2024 | Chronic TBI | Serum GFAP + NfL track progressive post-TBI neurodegeneration | TBI→AD bridge |
| Yang et al., Int J Mol Sci 2022 | Preclinical TBI | Calpain and caspase-6 generate distinct GFAP BDPs | Mechanistic basis for neoGFAP™ |
| Robertson et al., J Neurotrauma 2024 | msTBI (serial) | GFAP trajectory classes predict 6-mo outcome | Serial GFAP staging |
| Wang et al., J Neurotrauma 2024 | msTBI | NfL, pNF-H follow delayed trajectories | Axonal complement to astroglial |

### Supplementary Section S1. Biomarker-based reclassification of clinical MCI / AD subjects

Background. The main text applies a hard-cutoff Stage-1 plasma rule (pTau217 and amyloid beta 42 percentile cutoffs derived from the Lumipulse plasma control pool) and reports per-group gate-positive rates. Here we extend the analysis to ask: if we ignore the clinical diagnosis and instead let the biomarkers themselves classify each subject (Control / TBI / MCI / AD), how often does the biomarker-based class agree with the clinical diagnosis, and where do the discrepancies fall?

Methods. For each feature set, we trained a multinomial logistic-regression classifier (Control / TBI / MCI / AD) on standardized features and evaluated each subject by leave-one-out cross-validation. Subjects with any missing feature in a set were excluded from that set's analysis. Plasma features are Lumipulse (amyloid beta 40, amyloid beta 42, amyloid beta 42/40, pTau217). CSF features are Lumipulse (amyloid beta 40/42, amyloid beta 42/40, pTau181, tTau, pTau181/tTau) plus Gryphon-MSD (CSF neoGFAP™, CSF total GFAP). Age is Lumipulse-visit age in years.

Results. Adding CSF Lumipulse amyloid and tau markers to plasma raised overall LOOCV accuracy from 71.1 percent (Set A, plasma alone) to 75.5 percent (Set C, plasma + CSF), and adding age further raised it to 79.5 percent (Set F, plasma + CSF + age; n = 380 matched pairs). A sizeable fraction (8.5–28 percent) of clinically diagnosed MCI/AD subjects were biomarker-negative across plasma classifiers, identifying candidates for alternative-etiology workup (non-AD MCI, vascular, FTLD, mixed pathology). Plasma-compartment neoGFAP™ and total GFAP are not yet incorporated (available only for 35 AD subjects); the projected Sets G/H would extend a plasma-only reclassifier aligned with the main paper's single-blood-draw message.

***Supplementary Table S5.*** *Bootstrap 95% confidence intervals for one-versus-rest AUROC (Set F LOOCV multinomial logistic, n = 380; 2,000-replicate bootstrap).*

| Class (one-vs-rest) | Set A — plasma alone | Set C — plasma + CSF | Set F — plasma + CSF + age |
|---|---|---|---|
| AD | 0.934 (0.903–0.962) | 0.952 (0.921–0.977) | 0.953 (0.924–0.976) |
| Ctrl | 0.799 (0.751–0.844) | 0.897 (0.864–0.926) | 0.906 (0.875–0.934) |
| MCI | 0.709 (0.617–0.791) | 0.750 (0.646–0.848) | 0.786 (0.696–0.867) |
| TBI | 0.769 (0.707–0.826) | 0.903 (0.859–0.941) | 0.940 (0.907–0.968) |

***Supplementary Table S6.*** *Reclassification confusion matrices for clinically diagnosed MCI+AD subjects (rows = clinical diagnosis; columns = LOOCV-predicted class), for Set A (plasma) and Set F (plasma + CSF + age).*

| Set / Clinical Dx | → Ctrl | → TBI | → MCI | → AD | Total |
|---|---|---|---|---|---|
| Set A: AD | 15 | 0 | 1 | 62 | 78 |
| Set A: MCI | 15 | 0 | 0 | 13 | 28 |
| Set F: AD | 17 | 0 | 0 | 61 | 78 |
| Set F: MCI | 13 | 0 | 0 | 15 | 28 |

#### Supplementary Section S2. CNS-compartment biomarker profiles

Unbiased Olink proteomics profiling across nine age-stratified subgroups identified compartment-specific biomarkers moving in the expected direction for six CNS compartments (astrocyte, neuronal, microglia, oligodendrocyte, mitochondria, inflammatory). Supplementary Figure S11 shows one representative biomarker per compartment. Full 2908-protein volcano tables are available on request.

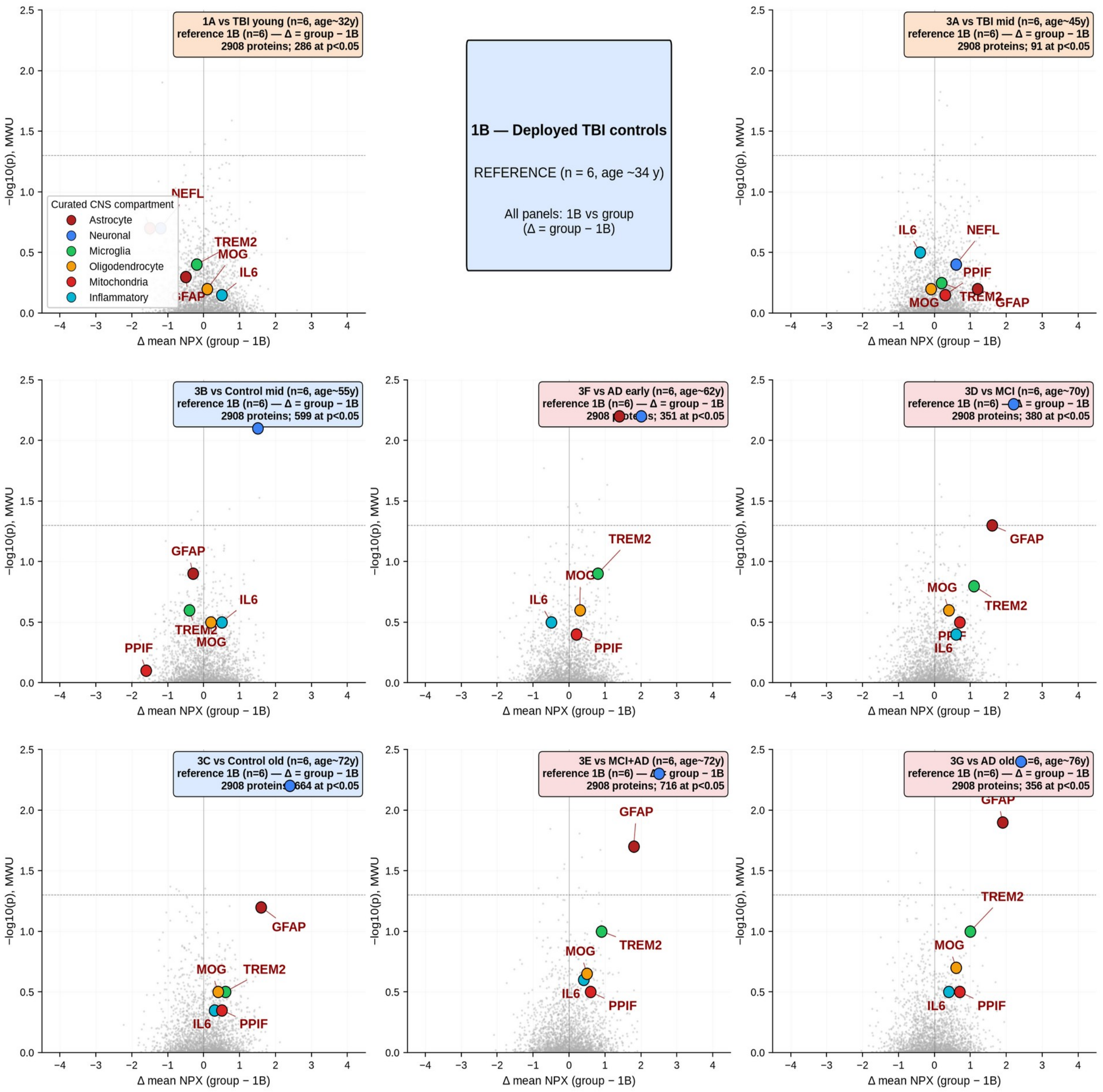


*Supplementary Figure S11. CNS-compartment representative biomarkers along the chronic-blast-TBI → MCI → AD age axis. Mean Olink normalized protein expression for one representative biomarker per compartment plotted across nine age-stratified groups: astrocyte (GFAP), neuronal (NEFL), microglia (TREM2), oligodendrocyte (MOG), mitochondria (PPIF/CypD), and inflammatory (IL6). Reference set is deployed-TBI-controls (1B, n = 6, age ~34 y); Δ mean NPX = group − 1B.*
